# Causal machine learning for assessing the effectiveness of off-label use of amiodarone in new-onset atrial fibrillation

**DOI:** 10.1101/2025.06.25.25330260

**Authors:** Simon Schallmoser, Jonas Schweisthal, Alexander von Ehr, Hamid Ghanbari, Fridtjof Schiefenhövel, Thomas S. Valley, Jenna Wiens, Stefan Feuerriegel

**Author notes:** Correspondence: Stefan Feuerriegel.

## Abstract

Off-label drug use, i.e., uses of a drug that differ from what regulatory authorities have approved, is common, occurring overall in up to 36% of prescriptions. Yet, the effectiveness across different patient subgroups is often poorly understood. In this study, we demonstrate how one can use causal machine learning (ML) together with real-world data to identify which patient groups are most likely to benefit from off-label use. Specifically, we assessed the effectiveness of off-label use of amiodarone in patients with new-onset atrial fibrillation (NOAF). NOAF can often lead to hemodynamic instability and rapid ventricular response, so that hemodynamic stability should be restored. We developed a causal ML model to predict individualized treatment effects (ITEs) of off-label amiodarone use on the probability of returning to hemodynamic stability. We used real-world data from the U.S. to develop the causal ML model and externally evaluated that model on real-world data from the Netherlands. Our predicted ITEs show that 44.8% (95% confidence interval [CI]: 38.4% to 51.0%) of patients benefit from off-label use of amiodarone with large heterogeneity: amiodarone is predicted to increase the probability of restoring hemodynamic stability by a mean of 0.5 percentage points (pp), with an interquartile range (IQR) of *−*1.1 pp to 1.0 pp, in the external dataset from the Netherlands. Using these ITEs, we defined a personalized treatment rule, which could increase the number of patients achieving hemodynamic stability by 4.4% (95% CI: 1.0% to 7.8%) compared to current practice. Additionally, we studied which biomarkers are predictive of treatment effect heterogeneity and found that patients with higher blood pressure may benefit most from off-label use of amiodarone. Altogether, our study shows the potential of causal ML together with real-world data in identifying patients who benefit from off-label drug use.

## Introduction

Off-label use of drugs refers to when a medication is administered under conditions for which it was not approved by regulatory authorities such as the U.S. Food and Drug Administration (FDA) [1]. Off-label use of drugs is widespread in clinical practice [2], occurring in 21% of drug orders overall [3]. It is especially common in intensive care units (ICUs) [4, 5, 6], where estimates from the U.S. suggest that around 36% of drug orders for adults in critical care are off-label [7].

Assessing the effectiveness of off-label drug use is challenging for several reasons. First, one often has to rely on real-world data such as electronic health records (EHRs) [8, 9, 10], as randomized controlled trials (RCTs) are absent or difficult to conduct, especially in ICUs [11]. Second, the effectiveness of off-label drugs may vary across individuals, benefiting some patients but not others [12, 13, 14, 15]. Hence, it is important to move beyond traditional, population-wide analyses and estimate personalized treatment effects in order to understand treatment effect heterogeneity. Third, there is a need to identify predictive biomarkers to inform new clinical guidelines but also support future research, such as confirmatory trials, and guide drug development. However, uncovering such predictive biomarkers, which explain treatment effect heterogeneity, is challenging because they must be recovered from patient data that is typically high-dimensional (e.g., EHRs encompass high-dimensional data including sociodemographic information, past treatments, prior diseases, and lab test results [16, 17]).

To overcome the above challenges, we propose the use of causal machine learning (ML) applied to routinely collected data to identify which patients are likely to benefit from off-label use. Causal ML is a branch of ML that develops flexible, data-driven methods for estimating treatment effects from data [18]. Causal ML is especially beneficial to our task. First, unlike standard predictive ML, causal ML aims at predicting the effect of an intervention, such as a drug, on a specific patient outcome from observational data [19, 20]. Second, causal ML is well-suited for making individualized predictions of treatment effects [21, 22, 23, 24] such as, for example, by computing individualized treatment effects (ITEs). This helps to capture heterogeneity in the treatment effectiveness across different patient profiles [17, 25, 26, 27]. Third, unlike conventional statistical approaches [28, 29], causal ML builds upon estimation strategies from machine learning [24] that offer efficient estimation in the presence of complex, high-dimensional datasets such as EHRs. Nonetheless, such causal ML approaches and the respective findings should be interpreted with appropriate caution and ideally validated through complementary analyses or prospective studies. This work illustrates how such approaches can be applied to challenging scenarios and used to generate hypotheses for further validation.

In this study, we develop a causal ML model to assess the effectiveness of off-label drug use of amiodarone in new-onset atrial fibrillation (NOAF). Patients with NOAF frequently experience a decline in hemodynamic stability, accompanied by rapid ventricular response [30]. NOAF with hemodynamic instability is a serious condition with increased risk of stroke, thromboembolism, and mortality [31]. Thus, the crucial objective is to restore hemodynamic stability, and, for this purpose, patients are frequently administered amiodarone, an antiarrhythmic agent [32]. Yet, amiodarone is only approved by the FDA for managing life-threatening ventricular arrhythmias, such as supraventricular tachycardia [33, 34], but not for NOAF where its use is off-label. In particular, for patients that experience hemodynamic worsening after NOAF, no clear guidelines exist on whether to use amiodarone [35, 36]. Previous studies have investigated the effectiveness of amiodarone [37, 38, 39, 40, 41, 42, 43, 44] but not with respect to restoring hemodynamic stability. Thus, it is unclear which patients with NOAF may benefit from amiodarone through improved hemodynamic stability.

Our causal ML model predicts the personalized treatment effects (i.e., ITEs) from off-label use of amiodarone on restoring hemodynamic stability in patients with NOAF, hemodynamic worsening, and rapid ventricular response. The predicted ITEs then show which patients benefit from off-label use and, therefore, for whom amiodarone thus leads to improved patient outcomes. We developed the causal ML model using real-world EHR data from the U.S. and externally evaluated the model on real-world EHR data from the Netherlands. Further, we calculated SHapley Additive exPlanations (SHAP) values [45] to understand the treatment effect heterogeneity and thus identify predictive biomarkers that can inform clinical guidelines. Finally, we used the predicted ITEs to develop a personalized treatment rule, which could be used to target treatment in patients who are most likely to benefit.

## Results

### Study populations

We performed a two-center study. To develop the causal ML model, we used the Medical Information Mart for Intensive Care IV (MIMIC-IV) dataset [46] from Physionet [47], a large dataset with electronic health records from patients that received critical care in the U.S. To further externally validate our causal ML model, we used the Amsterdam University Medical Centers Database (AUMCdb) [48], which provides a large-scale resource with EHRs from ICU patients from the Netherlands. For each, we then retrieved the stays from patients who experienced NOAF with a subsequent worsening of hemodynamic stability in combination with rapid ventricular response. Eventually, we obtained data from 705 stays of 494 patients (MIMIC-IV) and from 230 stays of 176 patients (AUMCdb). A flow chart showing the inclusion criteria is provided in Figure 1 (see Methods for more details).

**Figure 1:**
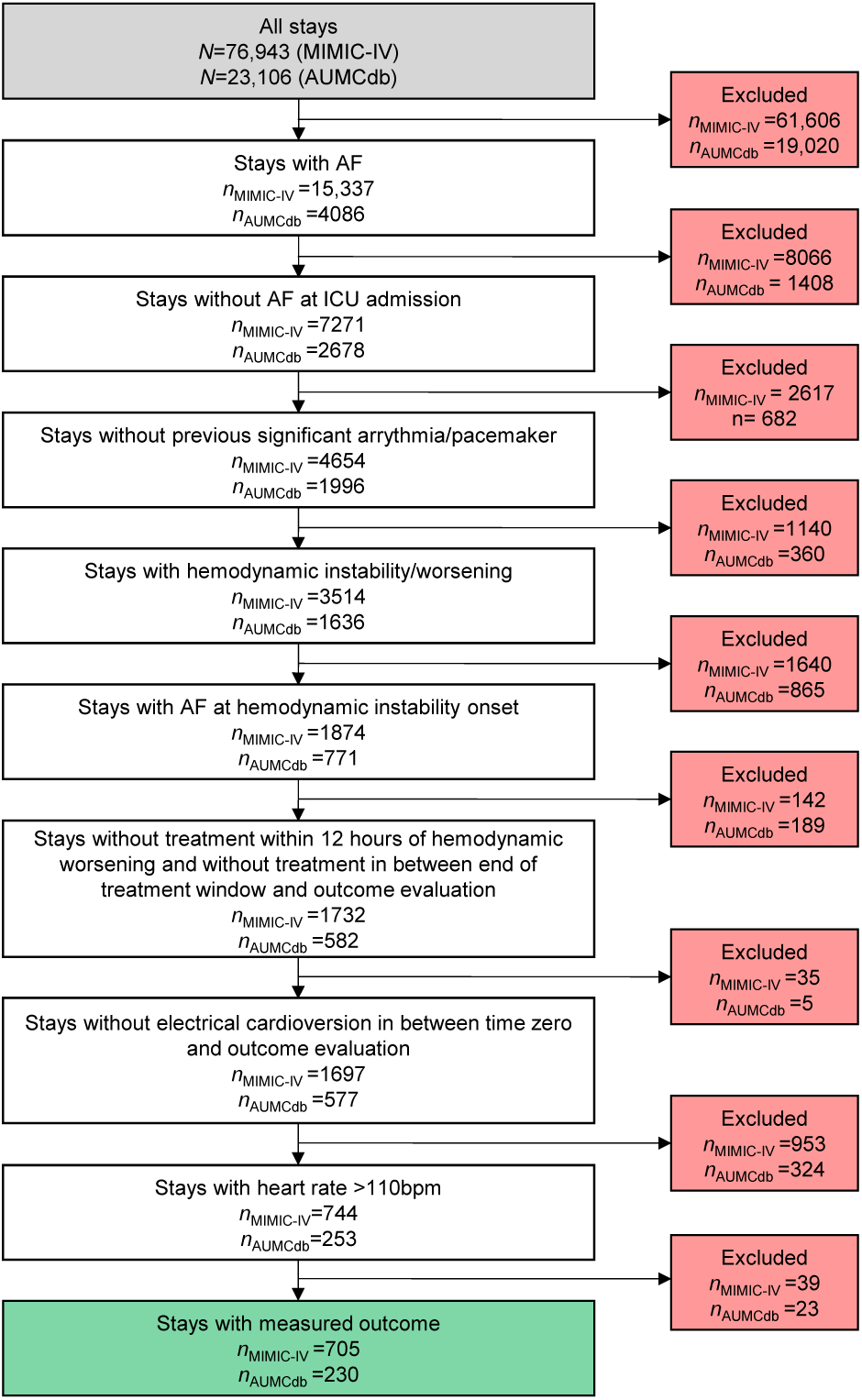
Flow chart of the inclusion criteria. The flow chart shows the inclusion criteria for the study population across both datasets. Reported are the numbers for the MIMIC-IV (*n*_MIMIC-IV_) and and AUMCdb datasets (*n*_AUMCdb_).

Amiodarone was administered in 96 out of 705 stays (13.6%) and in 73 of 230 stays (31.7%), in MIMIC-IV and AUMCdb respectively. Notably, the percentage of treated patients in AUMCdb is almost three times higher than in MIMIC-IV, which reflects differences in clinical practices. For example, amiodarone is generally used more frequently in AUMCdb than in MIMIC-IV, even for patients without NOAF.

The primary outcome in the subsequent analysis is hemodynamic stability (additionally, rhythm control and rate control are analyzed as further outcomes in Supplement D and Supplement E, respectively). Hemodynamic stability was restored in 15.6% of treated and 15.3% of control group patients in MIMIC-IV. Similarly, in AUMCdb, 16.4% of the treated and 19.1% of the control group patients achieved hemodynamic stability. Treated patients exhibited lower blood pressure, shorter AF duration, and higher heart rate compared to those in the control group (see Table 1). The time line for determining the inclusion criteria, observing off-label use of amiodarone, and assessing the patient outcomes is shown in Figure 2a

**Table 1:** Patient characteristics. Reported are the characteristics at the onset of hemodynamic worsening. The primary outcome in the analysis was hemodynamic stability. Additional outcomes,i.e., rhythm and rate control, are analyzed in Supplement D and Supplement E, respectively. Dichotomous variables are reported as counts (%), continuous variables as mean (SD). Abbreviations: ICU, intensive care unit; NOAF, new-onset atrial fibrillation; AF, atrial fibrillation; MAP, mean arterial pressure; SBP, systolic blood pressure; DBP, diastolic blood pressure; SD, standard deviation.

|  | MIMIC-IV (*n*=705) |  | AUMCdb (*n*=230) |  |
| --- | --- | --- | --- | --- |
|  | Treated | Control | Treated | Control |
| **Sample size** | 96 (13.6%) | 609 (86.4%) | 73 (31.7%) | 157 (68.3%) |
| **Outcomes** |  |  |  |  |
| Hemodynamic stability | 15 (15.6%) | 93 (15.3%) | 12 (16.4%) | 30 (19.1%) |
| Rhythm control | 56 (58.3%) | 311 (51.1%) | 40 (54.8%) | 84 (53.5%) |
| Rate control | 75 (78.1%) | 449 (73.7%) | 56 (76.7%) | 113 (72.0%) |
| **Baseline characteristics** |  |  |  |  |
| NOAF time [hours] | 46.4 (61.8) | 46.6 (80.9) | 65.8 (106.9) | 65.6 (103.7) |
| AF duration [hours] | 2.0 (4.1) | 6.7 (27.6) | 1.9 (2.1) | 9.2 (28.9) |
| Age [years] | 69.3 (10.0) | 71.7 (12.7) | 69.3 (13.7) | 70.2 (13.1) |
| Female | 44 (45.8%) | 308 (50.6%) | 31 (42.5%) | 66 (42.0%) |
| Heart rate [bpm] | 137.9 (17.2) | 128.2 (15.0) | 143.4 (24.5) | 133.6 (18.1) |
| MAP [mmHg] | 80.1 (15.6) | 83.4 (15.6) | 81.3 (17.1) | 84.5 (19.5) |
| SBP [mmHg] | 112.7 (22.0) | 116.4 (22.1) | 116.0 (29.1) | 124.9 (33.9) |
| DBP [mmHg] | 66.3 (15.7) | 70.0 (15.4) | 63.3 (14.3) | 65.2 (17.5) |
| Receiving inotropes | 26 (27.1%) | 123 (20.2%) | 44 (60.3%) | 77 (49.0%) |
| Mechanically ventilated | 35 (36.5%) | 203 (33.3%) | 43 (58.9%) | 100 (63.7%) |

**Figure 2:**
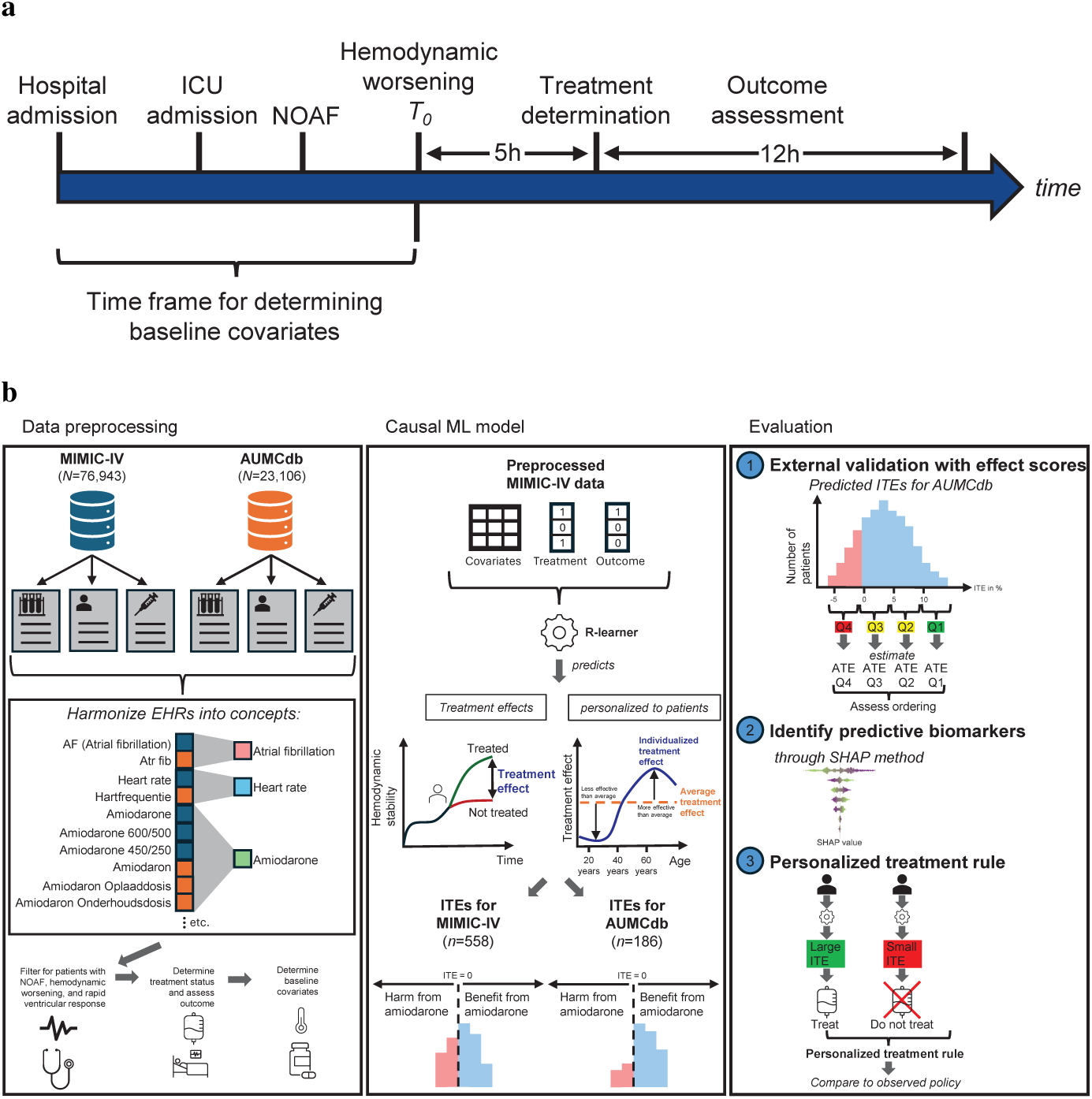
Workflow for personalized treatment effect predictions of off-label drug use. **a**, Time line for determining patient inclusion, off-label drug use, as well as for assessing patient outcomes and for recording the baseline covariates of patients. **b**, Development and evaluation of our causal ML model. First, the EHR data from MIMIC-IV and AUMCdb are preprocessed into harmonized concepts, which are used to select patients with NOAF, hemodynamic worsening, and rapid ventricular response. Further, for each patient, treatment and outcome are assessed and baseline predictors retrieved. Second, we used the preprocessed data from MIMIC-IV to train our causal ML model. The causal ML model can then be used to predict the ITEs and thus identify patients who benefit from off-label amiodarone use. Third, we externally validated our causal ML model in an independent cohort (AUMCdb). Further, we calculated SHAP values for MIMIC-IV and AUMCdb using our causal ML model to derive predictive biomarkers, i.e., covariates that are associated with a positive treatment response. Finally, we used the predicted ITEs to derive a personalized treatment rule and then compared the outcomes under our personalized rule to the outcomes under current practice.

### Predicted treatment effects

We developed a causal ML model to predict the ITEs of amiodarone on restoring hemodynamic stability (Figure 2b). Note that, when we refer to ITE in this paper, we mean individualized estimates of treatment effects characterized by some covariates that capture the patient profile. These should not be conflated with unobservable individual treatment effects [49]. Causal ML is a branch of ML techniques designed to model the relationship between an intervention and outcomes [19, 20]. In our case, we used causal ML to predict the patient outcomes (here: hemodynamic stability) in response to off-label use of amiodarone. Causal ML is particularly well-suited for our task because it enables individualized predictions of outcomes under interventions while accounting for the heterogeneity across different patient profiles captured in high-dimensional EHR datasets. The EHRs in this study contained information on socio-demographics, vital signs, laboratory measurements, medical procedures, medications, and administered fluids, which were all used as predictors.

To predict ITEs, we used the R-learner [50, 51], a state-of-the-art method for this purpose. The R-learner is a flexible and robust causal ML method, which uses a two-stage process (for other causal ML models see Supplement F). First, it removes confounding by estimating outcome and treatment models and then focuses on estimating the treatment effect, making it particularly well-suited for high-dimensional and heterogeneous data such as EHRs. See the Methods for further details.

The distribution of predicted ITEs for MIMIC-IV and AUMCdb are shown in Figure 3. The ITE is the difference in probability of restoring hemodynamic stability under amiodarone treatment versus no treatment. The average probability of returning to hemodynamic stability is 0.3 percentage points (pp) larger with amiodarone use than without for MIMIC-IV and 0.5 pp larger for AUMCdb, suggesting that amiodarone is sometimes effective. Yet, the corresponding interquartile ranges (IQRs) are [−1.4 pp, 1.2 pp] for MIMIC-IV and [−1.1 pp, 1.0 pp] for AUMCdb. This indicates that there is heterogeneity in the predicted ITEs: off-label use of amiodarone helps only some patients, but harms others. In MIMIC-IV, 45.0% of patients exhibited positive predicted ITEs, suggesting that they are likely to benefit from amiodarone in regaining hemodynamic stability. Further, 44.8% of patients in AUMCdb had positive ITEs. The majority of patients in both datasets experienced negative ITEs, meaning that for them the use of amiodarone would be harmful.

**Figure 3:**
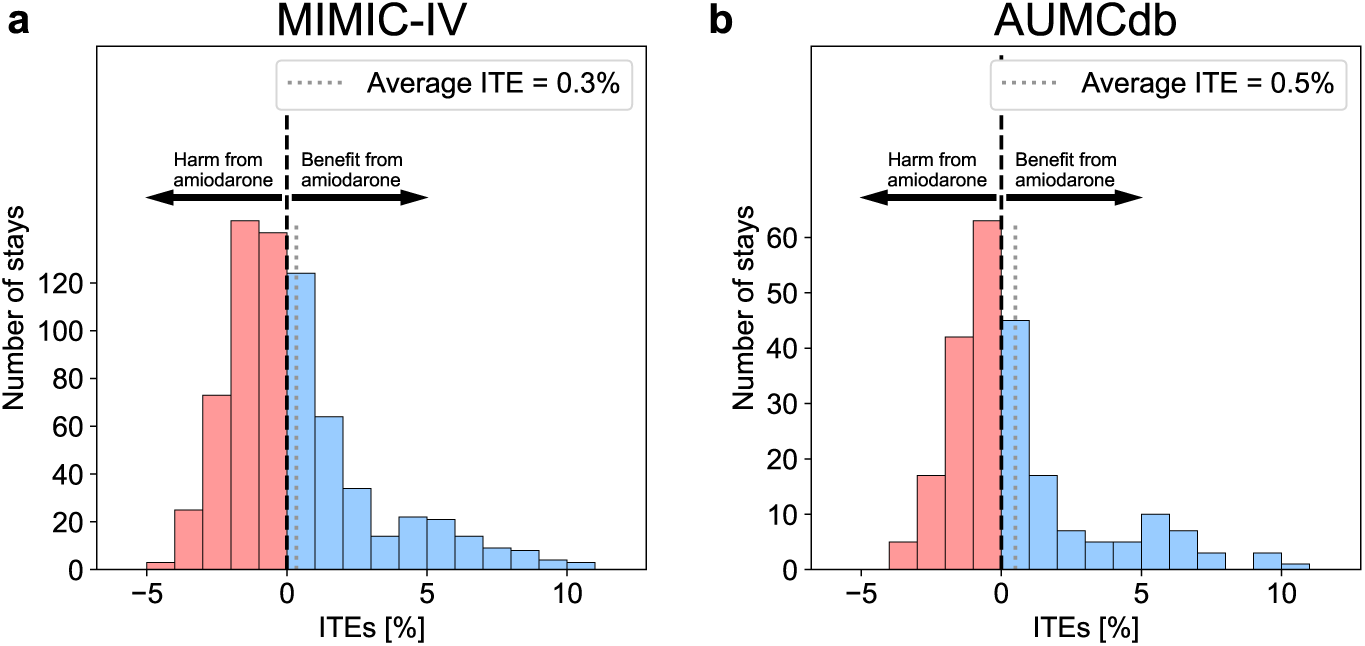
Heterogeneity of predicted ITEs across patients. The histograms show the distribution of predicted ITEs (in %) of amiodarone on restoring hemodynamic stability and their mean for **a**, MIMIC-IV and **b**, AUMCdb. The ITE is the difference in probability of restoring hemodynamic stability under amiodarone treatment versus no treatment. The distributions of ITEs show that heterogeneity in the effectiveness of amiodarone exists, with values ranging from -4.8% to 10.8% in MIMIC-IV and -4.0% to 10.0% in AUMCdb).

### Predictors of treatment effect heterogeneity

We now aim to identify covariates that explain treatment effect heterogeneity and thus present predictive biomarkers associated with a positive treatment response. For this, we calculated SHAP values [45], which quantify the contribution of each covariate to the variation in the treatment effects predicted by the causal ML model. For our analysis, SHAP provides a consistent approach to interpreting complex outputs of the causal ML model by attributing the influence of each covariate to the predicted ITE. Hence, positive SHAP values for a covariate indicate that the corresponding covariate is responsible for a more positive ITE, which contributes to a benefit from off-label use of amiodarone.

Figure 4 shows the 20 most relevant covariates influencing the predicted treatment effect heterogeneity. Our analysis suggests that amiodarone treatment may be particularly beneficial for patients with higher blood pressure since mean arterial pressure, systolic blood pressure, and the administration of antihypertensive agents are among the most important covariates. Further, older patients, as well as those with elevated red blood cell counts and reduced white blood cell counts, may also benefit more from amiodarone. Overall, the results across both datasets are largely consistent; i.e., 16 out of the top-20 covariates are the same. When comparing the importance across all predictors, we get a high correlation between both datasets (Pearson correlation coefficient was 0.946, *P <* 0.001).

**Figure 4:**
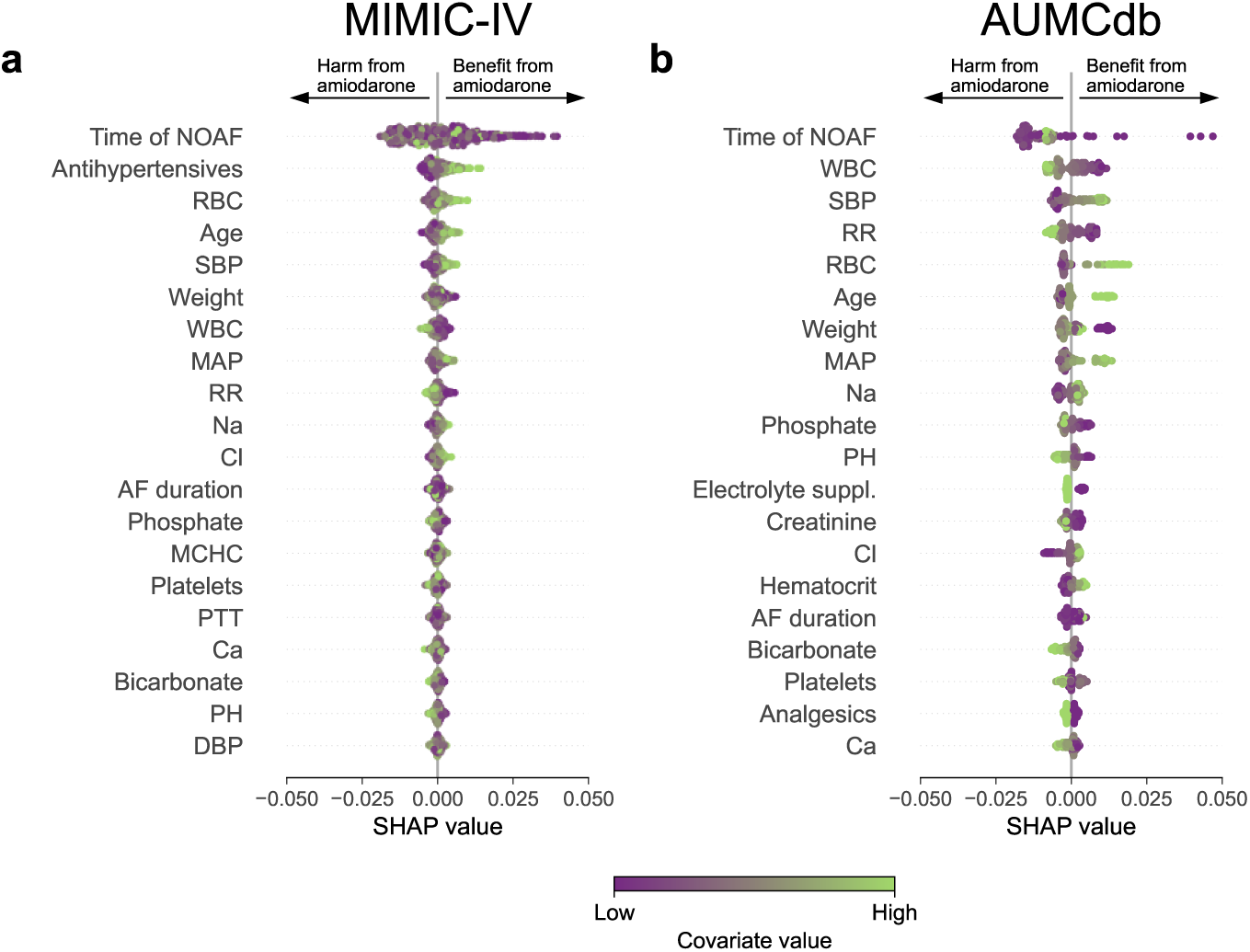
Predictors of treatment effect heterogeneity. The most relevant predictors of treatment effect heterogeneity for **a**, MIMIC-IV and **b**, AUMCdb. The analysis uses the SHAP values, which helps to explain the contribution of each covariate to the treatment effect predicted by the causal ML model. The predictors are sorted in descending order according to their relevance (top-to-bottom). Each dot in the visualization represents a single ICU stay. The color of the dot indicates the value of the corresponding covariate: green dots represent higher values of the covariate, while purple dots represent lower values. For example, patients with high values of red blood cell count are more likely to benefit from amiodarone than patients with low values. Abbreviations: RBC, red blood cell count; SBP, systolic blood pressure; WBC, white blood cell count; MAP, mean arterial pressure; RR, respiratory rate; Na, sodium; Cl, chloride; MCHC, mean corpuscular hemoglobin concentration; PTT, partial thromboplastin time; Ca, calcium; DBP, diastolic blood pressure.

To better understand how specific covariates related to the effectiveness of amiodarone, we analyzed the characteristics of patients who experienced treatment success compared to those who did not (Supplement H). Specifically, we stratified patients based on the predicted ITE and then compared the characteristics across patients with a positive versus negative ITE. This helps to characterize patients with treatment success. Similar to the SHAP analysis, patients with higher values of blood pressure (mean, systolic, and diastolic) were predicted to show benefit from amiodarone.

### Personalized treatment rule

The predicted ITEs can be used to derive a personalized treatment rule, which was defined as follows: treat the *x*% patients with the largest, predicted ITEs, where *x* is the proportion of actually treated patients in the dataset. The design of the personalized treatment rule has the advantage that we do not overstate the gain by simply treating more patients but rather that the gain arises from treating those patients that benefit most from amiodarone treatment.

Subsequently, the personalized treatment rule was evaluated by analyzing the improvement of patient outcomes compared to the observed treatment. Inspired by [17], we simulated the outcomes (i.e., whether a patient regains hemodynamic stability) under our personalized treatment rule and under current practice.

Using our personalized treatment rule, we estimate a 4.4% increase (95% CI, 1.0% to 7.8%) in the number of patients achieving hemodynamic stability compared to current practice.

Additionally, the personalized treatment rule can be evaluated by comparing two groups: (i) the group where the personalized treatment rule is concordant to the observed treatment and (ii) the group where it is discordant [16, 52]. In order to reduce the impact of confounding on our evaluation, we used a population that has been matched on the predicted propensity score (i.e., the predicted probability of receiving the treatment). The results are shown in Figure 5a. We find that 20.0% (95% CI, 9.7% to 30.3%) of patients regain hemodynamic stability, where the personalized treatment rule is concordant to the observed treatment. Contrary, only 15.1% (95% CI, 6.3% to 23.8%) of patients achieve hemodynamic stability, where the personalized treatment rule is discordant to the observed treatment. The average in the entire population of AUMCdb for regaining hemodynamic stability is 17.6%.

**Figure 5:**
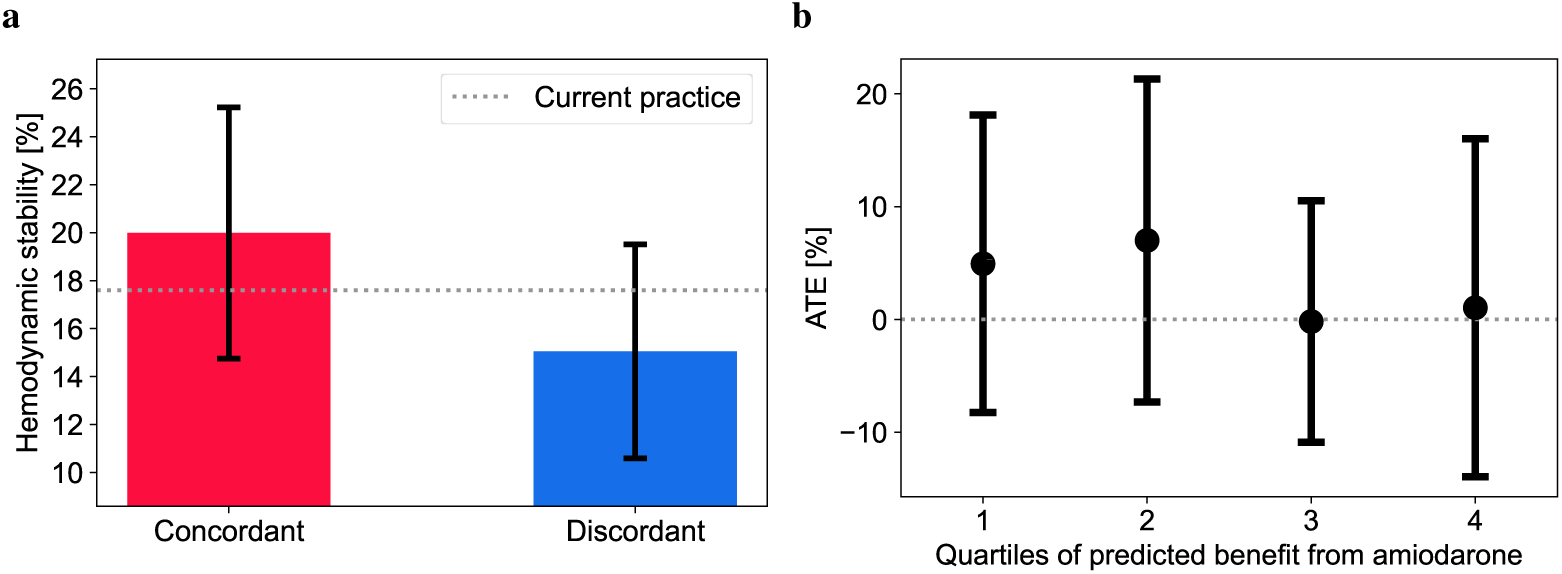
External validation of the causal ML model. **a**, A personalized treatment rule was derived using the predicted ITEs. Average patient outcomes (hemodynamic stability) were compared between patients where our personalized treatment rule was concordant with the observed treatment versus where it was discordant. The whiskers represent the standard errors (SEs), which was estimated using bootstrapping with 1,000 resamples. As a baseline, the average patient outcome under current practice is shown. **b**, The predicted ITEs for AUMCdb were sorted in descending order and split into quartiles. ATEs were estimated within each quartile, where we find that the two quartiles predicted to benefit most also showed the highest ATEs. This indicates that the causal ML model is robust. The whiskers represent the SEs.

### Model validation

To validate our model, we followed best practice in causal ML [15, 17, 19, 26]. First, we additionally estimated the ATE of amiodarone on restoring hemodynamic stability using an alternative approach (i.e., we applied the augmented inverse propensity weighting (AIPW) estimator [53]). Here, the estimated ATEs for MIMIC-IV and AUMCdb are 0.5% (95% confidence interval [CI]: −7.4% to 8.3%) and 2.6% (95% CI: −10.3% to 15.5%). For comparison, the averages over the ITEs are 0.3 (95% CI: −5.0% to 5.7%) and 0.5 (95% CI: −4.7% to 5.5%). Both the estimated ATE and the average ITE are similar, which demonstrates that the predictions are robust.

Further, we performed an external validation in an independent cohort (AUMCdb). Here, we use the effect scores method [15] to assess whether the predictions from our causal ML model are consistent [26, 17], that is, whether the ordering of our predicted ITEs in the independent cohort is consistent with those from other estimation techniques (Figure 5a). For this, we ranked patients based on their predicted ITEs for the AUMCdb dataset in descending order and divided them into four strata (quartiles). We then computed the ATE using the augmented inverse probability weighting (AIPW) estimator [53] for each stratum in the independent cohort. Hence, strata belonging to patients with a higher (lower) predicted ITE should also have a higher (lower) ATE. We find supporting results: the ATEs within the four quartiles (in descending order) are 4.9% (95% CI: −20.9% to 30.8%), 7.0% (95% CI: −21.0% to 35.1%), −0.2% (95% CI: −21.2% to 20.8%), and 1.0% (95% CI: −19.9% to 22.0%). Thus, our causal ML model can successfully identify patients who have a large benefit from off-label amiodarone use.

### Rhythm and rate control as outcomes

For patients with NOAF, hemodynamic worsening, and rapid ventricular response, other outcomes may be of interest such as achieving (a) rhythm control (i.e., restoring sinus rhythm) or (b) rate control (i.e., lowering the heart rate under a threshold of 110 bpm). Thus, we built causal ML models for rhythm and rate control as outcomes and repeated our above analyses. For rhythm control, 50.4% and 35.7% of predicted ITEs are positive for MIMIC-IV and AUMCdb, respectively and for rate control, 55.0% and 45.7% of predicted ITEs are positive. Details are in Supplement D (for rhythm) and Supplement E (for rate control).

## Discussion

Off-label drug use is prevalent in clinical practice [2, 3, 4, 5, 6], highlighting the need for approaches that generate robust clinical evidence and thus identify patient populations for which off-label drug use is both effective and safe. However, assessing the effectiveness of off-label drug use is inherently challenging, due to the absence of randomization in real-world data and the large variation in treatment responses across patients. In this study, we thus developed a causal ML model to assess the effectiveness of off-label use of amiodarone on restoring hemodynamic stability in ICU patients with NOAF, subsequent hemodynamic worsening, and rapid ventricular response. Understanding the effectiveness of amiodarone in this patient subgroup is highly relevant due to the lack of established guidelines [31, 35, 36]. Although amiodarone is frequently used in such cases [32], its use remains off-label. We found significant heterogeneity in treatment effectiveness. Using our causal ML model, we identified the subset of patients who benefit from amiodarone. Specifically, 45.0% of patients from the U.S. and 44.8% of patients from the Netherlands showed improved hemodynamic stability after amiodarone use. Notably, patients with higher blood pressure appeared to benefit the most.

This study offers real-world evidence about the effectiveness of off-label use of amiodarone for restoring hemodynamic stability. To the best of our knowledge, no prior research has addressed this specific question, particularly in the clinically-relevant subgroup of patients with NOAF followed by hemodynamic worsening and rapid ventricular response. In contrast, the effectiveness of amiodarone has only been studied with respect to other outcomes (e.g., rhythm and rate control) or treatments that are not applicable for patients with hemodynamic instability [37, 38, 39, 40, 41, 42, 43, 44]. Other research has analyzed which oral anticoagulant is most appropriate for each patient [54]. Yet, importantly, no prior study has attempted to identify specifically which patients benefit from amiodarone. This is crucial since NOAF in combination with hemodynamic instability and rapid ventricular response is a serious health condition and requires appropriate treatment. Further, amiodarone can have serious adverse effects, which limit broad application in clinical practice.

Our causal ML model has several strengths. First, we employed a state-of-the-art causal ML model, the R-learner [50, 51], to predict ITEs. This enables a data-driven path to developing personalized treatment rules. Second, we used real-world data to develop and evaluate our causal ML model. In particular, we used two large, independent ICU datasets from different countries with possibly different clinical routines and treatment strategies, to externally validate our results. Third, we identified predictive biomarkers linked to a positive treatment response. These predictive biomarkers can help characterize subgroups of patients that benefit from off-label use of amiodarone, thus providing the groundwork for more targeted clinical guidelines and future confirmatory RCTs.

Personalized treatment rules derived from our causal ML model can serve as a decision support tool for physicians in ICUs. Once a patient with NOAF develops hemodynamic worsening in combination with a rapid ventricular response, our causal ML model could be used to predict the patient-specific ITE, which is thus tailored to the unique clinical profile of the patient. This allows the treating physician to assess the predicted benefits of amiodarone treatment against the risks of potential adverse events For example, patients predicted to achieve hemodynamic stability without amiodarone could avoid unnecessary exposure to the drug and its associated side effects, thus minimizing potential harm. Conversely, patients unlikely to benefit from amiodarone, alternative interventions, such as immediate electrical cardioversion, could be prioritized to optimize outcomes. By enabling personalized treatment strategies, this approach could reduce the burden of adverse effects associated with amiodarone and generate clinical evidence to develop guidelines tailored to individual patient profiles. Nevertheless, we recommend a cautious application of the model in clinical practice. A necessary next step is to design and validate the model in a confirmatory RCT to establish the efficacy and safety in guiding treatment decisions.

Our study has limitations, which are inherent to the use of real-world data and the retrospective nature of our analysis. First, predicting treatment effects from EHR data relies upon assumptions, such as the absence of unobserved confounding, which are difficult or even impossible to test. We mitigated this risk by including a large number of clinical, demographic, and physiological covariates (74 in total) to generate confounder-adjusted predictions, so that the likelihood of unobserved confounding is limited. Second, patients with missing outcome data, such as those who died or were discharged before hemodynamic stability could be assessed, were excluded from the analysis. However, this corresponds to less than four percent of the patients in our sample, because of which the impact on our analysis is minimal. Third, validating predictions of treatment effects is notoriously challenging. We thus addressed this by following best practice in causal ML [16, 17, 19, 26, 52]. In particular, we externally validated our causal ML model in an independent cohort, which demonstrates that the predicted ITEs by our causal ML model are robust across datasets and corroborates the generalizability of our results. Hence, one of the next steps is to design an RCT to validate the effect of our causal ML model in clinical practice and thus show the clinical benefit from optimizing off-label treatment strategies.

In sum, our study highlights the potential of causal ML combined with real-world data to evaluate the safety and effectiveness of off-label drug use in critical care. By identifying patient subgroups who are most likely to benefit, our framework can generate new clinical evidence and support the development of personalized guidelines for off-label treatments. The framework is flexible and can be applied to other drugs that are used off-label.

## Methods

### Study design

This study was designed as a retrospective cohort study, where we aimed to develop a causal ML model to predict the ITEs of amiodarone on restoring hemodynamic stability for patients with NOAF, hemodynamic worsening, and rapid ventricular response in the ICU. For this, we used two distinct datasets, one for the development of the causal ML model and the other for external evaluation. To develop our causal ML model, we used the MIMIC-IV dataset (version 2.0) [46, 47], which is a database containing EHRs from the Beth Israel Deaconess Medical Center in Boston, Massachusetts, USA. MIMIC-IV includes data from 53,569 patients (76,943 stays) admitted to the ICU between 2008 and 2019. To externally assess the robustness of the causal ML model, we chose the AUMCdb dataset (version 1.0.2) [48]. AUMCdb contains data from 20,109 patients (23,106 stays) admitted to the ICU or medium care unit (high-dependency unit) of the Amsterdam University Medical Center in the Netherlands between 2003 and 2016. Since only de-identified retrospective data were used, no ethical approval was needed by LMU Munich.

### Inclusion criteria

In this study, patients with NOAF were included, when hemodynamic instability occurred subsequent to NOAF. For patients who were already hemodynamically unstable at the onset of NOAF, only those in whom NOAF led to a further worsening of hemodynamic instability were included (see definition below). To identify patients with NOAF, we used heart rhythm annotations that were recorded at the bedside. NOAF was defined as a measurement of AF without AF as the first heart rhythm annotation after ICU admission to ensure that no patients with persistent or permanent AF were included. Further, we excluded patients with pacemakers or previous, serious arrhythmias such as atrial flutter, (supra-)ventricular tachycardia, and third-degree atrioventricular block. Since heart rhythm measurements were only charted roughly hourly, we set the time of NOAF to the time point at which the last heart rhythm measurement was recorded prior to the first AF measurement.

Hemodynamic stability was assessed using mean arterial pressure (MAP) measurements and administration records of inotropes. Patients were considered hemodynamically unstable if they fulfilled one of the following conditions: MAP less than 80 mmHg, receiving norepinephrine with a rate larger or equal to 0.1 µg/kg/min, or receiving other inotropes (epinephrine, dopamine, dobutamine, or milrinone) regardless of the rate. For hemodynamically stable patients at NOAF, patients were only included if they developed hemodynamic instability following NOAF. For hemodynamically unstable patients at NOAF, patients were included if hemodynamic instability worsened following NOAF. A worsening of hemodynamic instability was defined as a relative decrease of MAP of more than 20% or a relative increase of inotrope rate of more than 20%. Time zero was defined as the time point at which hemodynamic instability occurred for previously hemodynamically stable patients. For hemodynamically unstable patients at NOAF, time zero was defined as the time point at which hemodynamic instability worsened. Finally, only patients with rapid ventricular response (i.e., an increased heart rate *>* 110 bpm at time zero) were included. Similar to heart rhythm measurements, MAP was typically recorded every hour; hence, MAP measurements were adjusted by shifting them backward to the prior measurement.

Patients were excluded if AF had already resolved at time zero. Patients were excluded from the control group if treatment was administered in the twelve hours before time zero and in between the end of the treatment window (i.e., five hours after time zero) and the end of the outcome horizon (i.e., seventeen hours after time zero, see Figure 2a). Further, patients were excluded if they received electrical cardioversion during the treatment window or outcome window. Additionally, we did not include patients for whom the outcome could not be assessed either due to death or discharge from the ICU before hemodynamic stability could be assessed (see Supplement G for a survival analysis including all patients).

### Treatment and outcome definitions

The treatment was defined as a dichotomous variable referring to an intravenously administered amiodarone bolus of at least 150 mg over a maximum period of 30 minutes. Patients were considered to be treated if the treatment was administered within a window of five hours after time zero; and untreated if the treatment was not administered in the time frame between time zero and the end of the outcome window.

The outcome of interest was hemodynamic stability, which can be influenced by amiodarone either by returning a patient’s heart rhythm to sinus rhythm (referred to as rhythm control) or by lowering a patient’s heart rate (here below 110 bpm, referred to as rate control). The outcome was assessed in a twelve hour window after the treatment window ended (i.e., in the five to seventeen hours after time zero). The outcome was defined as a dichotomous variable denoting whether the patient regained hemodynamic stability in combination with either rhythm or rate control for at least two thirds of the time in the outcome window. A patient was considered to have regained hemodynamic stability if (i) MAP was higher or equal to 80 mmHg or higher than the baseline value that defined a patient hemodynamically unstable at NOAF and (ii) no inotropic agents were intravenously administered or the rate of inotropic agent administration was smaller or equal to the baseline rate that defined a patient hemodynamically unstable at NOAF.

### Covariates

We included a variety of baseline covariates that may predict ITEs and that may act as confounders [55, 56], that is, covariates that influence both the treatment decision of the physician and the outcome. For example, we included vital measurements such as heart rate and blood pressure, laboratory measurements such as hemoglobin and creatinine, medications such as norepinephrine and beta blockers, and fluids such as 0.9% sodium chloride and Ringer’s lactate solution. For vital and laboratory measurements, we included the latest measurement prior to time zero. Covariates pertaining to vital and laboratory measurements with more than 25% missing values were excluded from the model. A detailed list of all 95 initially considered covariates, as well as the final 74 covariates, is provided in Supplement A.

We performed the following preprocessing steps. Categorical covariates were one-hot encoded. Missing data of continuous and dichotomous covariates were imputed using the median and mode, respectively. Finally, covariates were standardized by subtracting the mean and dividing by the standard deviation.

### Causal ML model

Causal ML refers to a set of methods that aim at predicting treatment effects [19]. Unlike standard, predictive ML, which aims at predicting outcomes, causal ML seeks to predict how interventions or changes in one variable will impact a certain outcome. The use of standard, predictive ML does not account for confounding variables and would thus lead to biased estimates [57]. Additionally, causal ML has the advantage over traditional statistics in that it can handle high-dimensional datasets such as EHRs as in our study. Further, causal ML can be used in combination with non-parametric ML models to capture non-linear relationships between covariates and treatment effects, thus enhancing the ability to personalize treatments for patients. We followed best practice for the causal ML workflow [19, 20].

In this study, we aim to predict individualized treatment effects (ITEs). The ITE quantifies the treatment effect within specific subgroups, thus accounting for variations in patient characteristics. By examining ITEs, we gain insights into not only the general effectiveness of amiodarone but also how its impact may differ for different patients, thus enabling a more personalized treatment strategy.

We applied the R-learner to predict ITEs [50, 51]. The R-learner is a flexible and robust causal ML approach designed for estimating treatment effects. It relies on two distinct ML models that are trained to predict (i) the treatment assignment (of amiodarone) and (ii) the outcome (hemodynamic stability). For both models, we used random forests, which are a non-parametric ML model that consists of multiple decision trees to obtain reliable estimates [58]. Using both models, estimates of the treatment as well as the outcome can be obtained, which are then subtracted from the true treatment and outcome information, resulting in the corresponding residuals. These residuals are then regressed on the covariates in a final stage ML model to obtain the ITEs. For this final stage regression model, we used a causal forest, which is an extension of the random forest that specifically aims at estimating treatment heterogeneity [22, 59]. To compute the ATE, we used the AIPW estimator [53]. Similar to the R-learner, it uses two ML models for predicting the treatment assignment and the outcome. Predictions of these two ML models are then combined to give a reliable prediction for the ATE, meaning that only one of the two models needs to be accurate to provide a reliable result. For these models, we again used random forests.

We used the *EconML* package to implement our causal ML model [60]. Further details of our causal ML model are provided in Supplement B.

### Personalized treatment rule

Inspired by [17], we build a personalized treatment rule using our predicted ITEs, which was defined as follows: “treat the *x*% patients with the largest, predicted ITEs, where *x* is the proportion of actually treated patients in the dataset”. Then, we simulated the outcomes (i.e., whether a patient regains hemodynamic stability) under our personalized treatment rule and under current practice. To this end, we trained two random forest classifiers [58], one for the treatment group and one for the control group, to predict whether a patient returns to hemodynamic stability. Using these predictions, we assessed whether more patients are expected to return to hemodynamic stability under our personalized treatment rule compared to current practice.

Further, we compared the average outcome (hemodynamic stability) of those patients, where the observed treatment is identical to the suggested treatment according to our personalized treatment rule (concordant group), to those patients, where both differ (discordant group). We evaluated this on a population that has been matched on the predicted propensity score (i.e., the predicted probability of receiving the treatment) to reduce the impact of confounding. If the average outcome within the concordant group is larger than in the discordant group, it suggests that our predicted ITEs are meaningful.

### Model validation and robustness

Since treatment effects are never observed, we cannot rely upon standard validation procedures from traditional, predictive ML [19]. Instead, we followed best practice in causal ML to validate our model [15, 17, 19, 26].

First, we compared the average of predicted ITEs against the ATE. If the ITE predictions are accurate, their average should approximate the ATE, indicating that the model captures meaningful heterogeneity in treatment effects.

Second, we assessed the ordering of our predicted ITEs (trained on MIMIC-IV) for an independent cohort (AUMCdb) to check whether the ordering is consistent with other estimation strategies. To this end, patients in AUMCdb were ranked according to their predicted ITEs in descending order and divided into four strata (quartiles). Then, we calculated the ATE using the AIPW estimator within each stratum, which has been trained on AUMCdb data. If strata belonging to patients with higher (lower) ITEs also show higher (lower) ATEs, there is reason to expect that our causal ML model for predicting ITEs is robust.

Third, we applied two alternative ITE estimators and compared their predicted ITEs with those obtained from our causal ML model. Our analysis revealed a strong correlation between the predicted ITEs, thereby underscoring the consistency and robustness of the predicted ITEs of our causal ML model. For more details, see Supplement F.

Fourth, we used an alternative estimation strategy, namely, a causal survival forest [61] to account for the time-to-event nature of our setting. For more details, see Supplement G.

Fifth, we applied three different refutation checks to assess the robustness of our causal ML model, i.e., we repeated the analyses with a random treatment, a random outcome, and by adding random covariates [62]. In the first two cases, the ITEs should disappear and in the last case, the ITEs should remain robust. For more details, see Supplement I.

Sixth, we assessed the calibration of the two ML models (one for the treatment assignment and one for the outcome) required for predicting ITEs with the R-learner. For more details, see Supplement J.

### Predictors of treatment effect heterogeneity

To identify the most relevant covariates of treatment heterogeneity, we calculated SHAP values [45]. SHAP offers an interpretable way to quantify each covariate’s contribution to the ITE, which was predicted by the causal ML model. The SHAP values were examined for the final stage model, i.e., the causal forest. Thus, we aimed to identify predictive biomarkers, covariates that predict individual variation in response to amiodarone. We used bee swarm plots to show not only the ranking of the most important predictors of treatment heterogeneity but also their direction, i.e., whether larger or smaller values of a covariate are associated with a larger ITE.

### Additional information

#### Code availability

Code that supports the findings of our study is uploaded as a supplementary file. The code will be made publicly available via a GitHub repository upon acceptance.

#### Data availability

The patient data used in this study are available via the Medical Information Mart for Intensive Care IV (MIMIC-IV) dataset [46, 47] and Amsterdam University Medical Centers Database (AUMCdb) [48].

### Author information

#### Author contributions

S.S. performed the data analysis. All authors contributed to conceptualization and results interpretation. S.S. and S.F. contributed to manuscript writing. All authors approved the manuscript.

#### Competing interests

The authors declare no competing interests.

## Acknowledgements

S.F. acknowledges funding via the Swiss National Science Foundation (SNSF), Grant 186932. The funding body had no control for the design, analysis, or interpretation of the study.

## Supplements

### Supplement A List of covariates

In order to obtain reliable estimates of ITEs using observational data, it is crucial to include all covariates that can influence both the treatment decision and the outcome. Here, we largely followed the work by Bedford et al. [55], in which a panel of experts in cardiology and intensive care medicine identified relevant covariates. We grouped the covariates into the following five categories: socio-demographic, vital signs, laboratory measurements, medical procedures, medications, and fluids. Covariates pertaining to vital and laboratory measurements with more than 25% missing values were not included.

Socio-demographic covariates include variables such as sex, age (at ICU admission), weight, height, time of NOAF, onset of hemodynamic worsening relative to NOAF, and the type of ICU, where we distinguished between medical, surgical, cardiac medical, cardiac surgical, and other. Vital signs are variables that are measured at the bedside such as heart rate and blood pressure. Laboratory measurements include variables such as white blood cell count and creatinine. For vital signs and laboratory measurements, we used the latest measurement prior to time zero (onset of hemodynamic worsening). Medical procedures consist of variables such as mechanical ventilation and hemofiltration. The medications consist of two different types of variables. First, we included rates of medications, where we are interested in the value exactly at time zero (e.g., norepinephrine rate). Medications of the second type are defined as dichotomous variables and denote whether medications were prescribed within the last 24 hours prior to time zero. For fluids, we assessed the amount administered in the 4 hours prior to time zero.

In MIMIC-IV, ICD-10 codes are used to describe comorbidities, which were recorded at the end of each hospital stay. Hence, we cannot use them as covariates since the exact onset of the comorbidity remains unclear. In AUMCdb, no information regarding comorbidities is recorded. Therefore, we used laboratory measurements and medication records to account for the comorbidities included in the list of covariates by Bedford et al. [55].

**Table S1:**
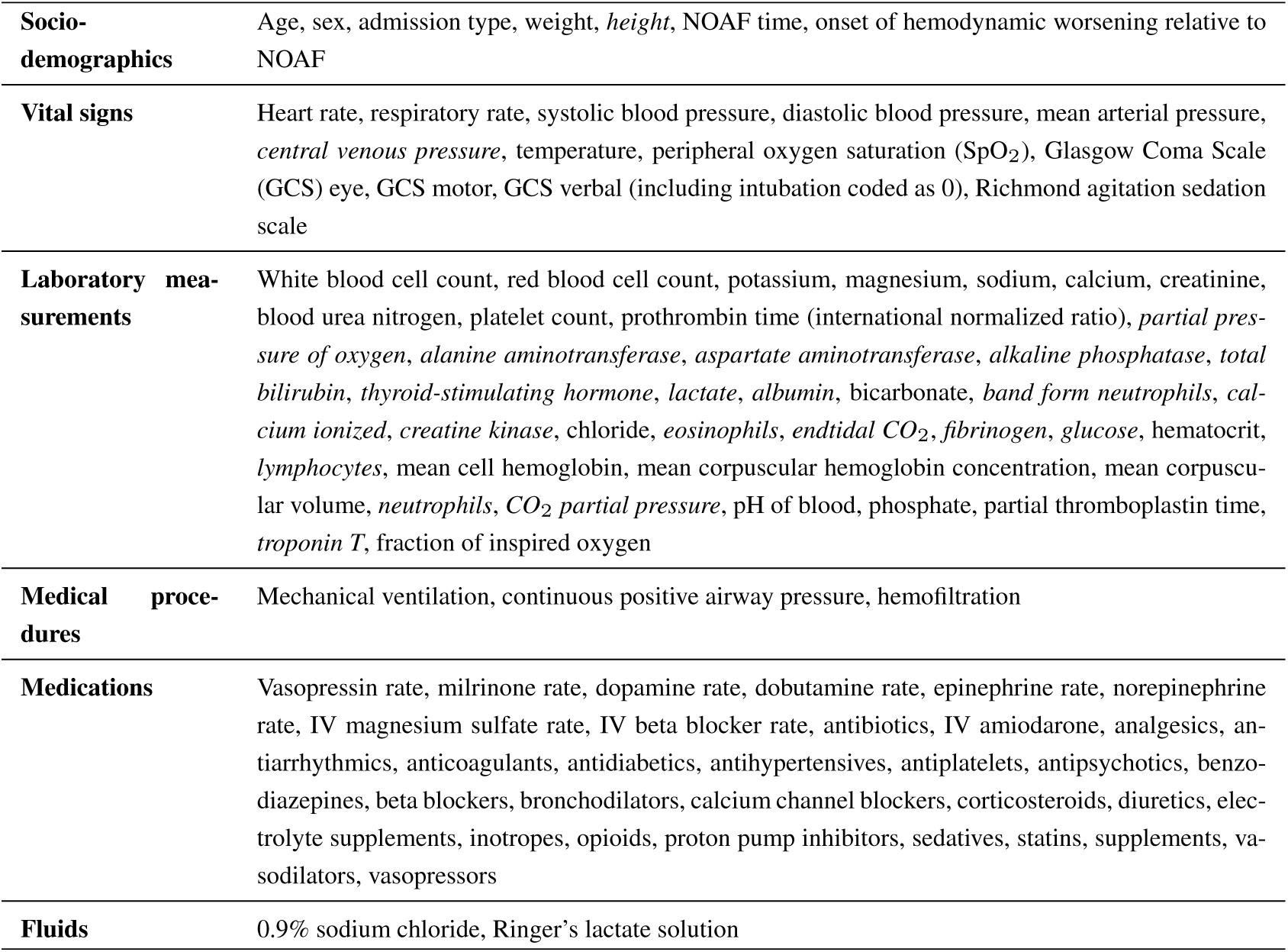
List of covariates. Covariates that were ultimately excluded due to too many missing values are highlighted in italics.

|  |  |
| --- | --- |
| **Socio-demographics** | Age, sex, admission type, weight, *height*, NOAF time, onset of hemodynamic worsening relative to NOAF |
| **Vital signs** | Heart rate, respiratory rate, systolic blood pressure, diastolic blood pressure, mean arterial pressure, *central venous pressure*, temperature, peripheral oxygen saturation (SpO<sub>2</sub>), Glasgow Coma Scale (GCS) eye, GCS motor, GCS verbal (including intubation coded as 0), Richmond agitation sedation scale |
| **Laboratory measurements** | White blood cell count, red blood cell count, potassium, magnesium, sodium, calcium, creatinine, blood urea nitrogen, platelet count, prothrombin time (international normalized ratio), *partial pressure of oxygen*, *alanine aminotransferase*, *aspartate aminotransferase*, *alkaline phosphatase*, *total bilirubin*, *thyroid-stimulating hormone*, *lactate*, *albumin*, bicarbonate, *band form neutrophils*, *calcium ionized*, *creatine kinase*, chloride, *eosinophils*, *endtidal CO<sub>2</sub>*, *fibrinogen*, *glucose*, hematocrit, *lymphocytes*, mean cell hemoglobin, mean corpuscular hemoglobin concentration, mean corpuscular volume, *neutrophils*, *CO<sub>2</sub> partial pressure*, pH of blood, phosphate, partial thromboplastin time, *troponin T*, fraction of inspired oxygen |
| **Medical procedures** | Mechanical ventilation, continuous positive airway pressure, hemofiltration |
| **Medications** | Vasopressin rate, milrinone rate, dopamine rate, dobutamine rate, epinephrine rate, norepinephrine rate, IV magnesium sulfate rate, IV beta blocker rate, antibiotics, IV amiodarone, analgesics, antiarrhythmics, anticoagulants, antidiabetics, antihypertensives, antiplatelets, antipsychotics, benzodiazepines, beta blockers, bronchodilators, calcium channel blockers, corticosteroids, diuretics, electrolyte supplements, inotropes, opioids, proton pump inhibitors, sedatives, statins, supplements, vasodilators, vasopressors |
| **Fluids** | 0.9% sodium chloride, Ringer's lactate solution |

### Supplement B Details on the causal machine learning model

In the following, we provide additional technical and implementation details of our causal ML model. For a detailed introduction to causal ML, we refer to the recent overviews in medicine [19, 20].

#### B.1 Technical details

We employ the potential outcomes framework [28] to define our causal quantities of interest, namely, the average treatment effect (ATE) and the individualized treatment effect (ITE). The potential outcomes framework provides a formal basis for understanding causal effects by comparing what happens to an individual under different treatment conditions. In this framework, each individual has different potential outcomes, namely, for each possible treatment, but only one of these outcomes is observed (i.e., the one corresponding to the treatment the individual actually received).

To estimate treatment effects using observational data, the following assumptions must be satisfied [28, 63]: (i) stable unit treatment value assumption (SUTVA), (ii) positivity, and (iii) unconfoundedness. Assumptions (i) and (iii) are inherently untestable, whereas assumption (ii) can be validated (see Supplement C). It is reasonable to expect that the assumptions are fulfilled in our study. (i) SUTVA implies that the data are independent and identically distributed and that consistency holds. Consistency states that the observed outcome for a given patient under a particular treatment is the same as the potential outcome that would have occurred had they been assigned to that treatment. This is likely to hold given how measurements are recorded and how treatments are administered in ICUs. (ii) Positivity, also referred to as overlap assumption, states that every patient has a non-zero probability of receiving each treatment option. It is reasonable to assume that the assumption is ensured as there are no evidence-based guidelines for amiodarone use in our patient population. Further, we manually inspected the propensity scores (see Supplement C), finding evidence that the assumption was fulfilled. (iii) Unconfoundedness implies that no unmeasured confounders exist, meaning that conditioned on all observed covariates, the treatment assignment is independent of the potential outcome. This is reasonable to expect given that we use a large number of clinical, demographic, and physiological covariates (74 in total), so that the risk of unobserved confounding is minimal. Further, we employed refutation checks [62] and a negative control outcome analysis [64] in order to test for unobserved confounding. However, the refutation checks suggest that unmeasured confounding was unlikely to bias the results.

The previous assumptions are required, regardless of whether ML or classical statistics is employed as an estimation strategy, due to the fundamental problem of causal inference [28, 65]. The fundamental problem of causal inference refers to the fact that we only observe the outcome for the treatment decision that has been made but not for the opposite treatment decision. Therefore, true treatment effects are never observed, because of which assumptions are needed for valid inference [28, 63]. This is also the reason why a standard, predictive ML model for predicting patient outcomes under treatment would be biased and thus unreliable [19].

Technically, the potential outcome for an individual *i* under treatment *T* = *t* is denoted by *Y_i_*(*t*), where *Y_i_*(*t*) represents the outcome that would be observed if individual *i* were assigned to treatment *t*. Then, we have *Y_i_*(1) as the potential outcome for individual *i* if they receive amiodarone (*T_i_* = 1) and *Y_i_*(0) as the potential outcome for individual *i* if they do not receive amiodarone (*T_i_* = 0).

We can then formally define the ATE as

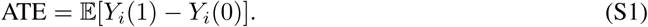

The ITE is further defined as

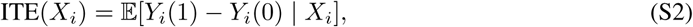

where *X_i_* denote the covariates of individual *i*.

To calculate the ATE, we applied the augmented inverse propensity weighting (AIPW) estimator [53]. For this method, it is necessary to train the following three prediction models: (i) for the treatment assignment (propensity score) defined as *e*^(*X_i_*) = P(*T_i_* = 1 | *X_i_*), (ii) for the outcome of the untreated individuals defined as *m*^ _0_(*X_i_*) = E[*Y_i_* | *T_i_* = 0*, X_i_*], and (iii) for the outcome of the treated individuals defined as *m*^ _1_(*X_i_*) = E[*Y_i_* | *T_i_* = 1*, X_i_*]. Using the three prediction models, the ATE can be calculated using the AIPW estimator via

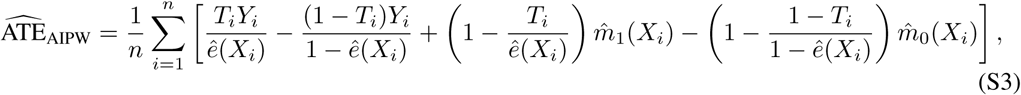

where *n* refers to the number of individuals.

To predict the ITE, we used the R-learner [50, 51], which involves the following prediction models: (i) for the treatment assignment (propensity score) defined as *e*^(*X_i_*) = P(*T_i_* = 1 | *X_i_*) and (ii) for the outcome defined as *m*^ (*X_i_*) = E[*Y_i_* | *X_i_*].

Using both prediction models, we then employ another final stage ML model to optimize the following objective function to obtain predictions for the ITE:

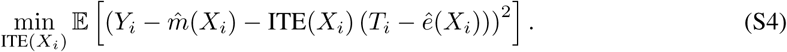

Here, it is crucial that the prediction models for treatment assignment and outcome, as well as the final prediction model for the ITE, are trained via cross-fitting [21, 50]. This involves splitting the data into two partitions: the first partition of the data is used to train the treatment and outcome models, which are then used to predict the treatment and outcome on the second partition of the data. The process is repeated with the roles of the two partitions reversed, thus obtaining estimates for the entire dataset.

For binary outcomes (as in our setting), the expectation operator (E) in the outcome models *m*(*X_i_*) for the ATE and the ITE may be replaced with a probability (P). In Supplement F, we compared the ITE predictions of the R-learner to those of two other causal ML models, namely a DR-learner and causal forest.

#### B.2 Implementation details

We followed best-practice and trained all models using 10 different random seeds in order to improve stability [27]. All predictions from these 10 models were averaged.

For all prediction models for the treatment and the outcome, we used random forests [58] based on the implementation in *scikit-learn* (version 1.2.2) [66]. We used 3-fold cross-validation to optimize the hyperparameters of these models by applying a random search with 10 iterations and the hyperparameter grid given in Table S2.

**Table S2:**
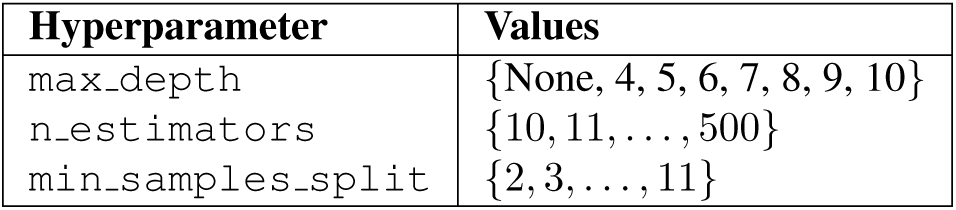
Hyperparameter grid for random forests.

| Hyperparameter | Values |
| --- | --- |
| max_depth | {None, 4, 5, 6, 7, 8, 9, 10} |
| n_estimators | {10, 11, ..., 500} |
| min_samples_split | {2, 3, ..., 11} |

We used the implementation of the R-learner from *EconML* (version 0.15.1) [60]. For the final stage ML model, we applied causal forests [22, 59] with the following hyperparameters as recommended by the developers of *EconML*, see Table S3.

To obtain ITE predictions and SHAP values for the training dataset (MIMIC-IV), we applied 20-fold cross-validation. The cross-validation is performed such that the splits are determined on the patient level, i.e., to prevent that one stay of a patient is used for training and another one for assessing the robustness. Afterward, we retrain the R-learner on the entire MIMIC-IV data and use this model to obtain predictions of the ITEs for AUMCdb patients.

**Table S3:**
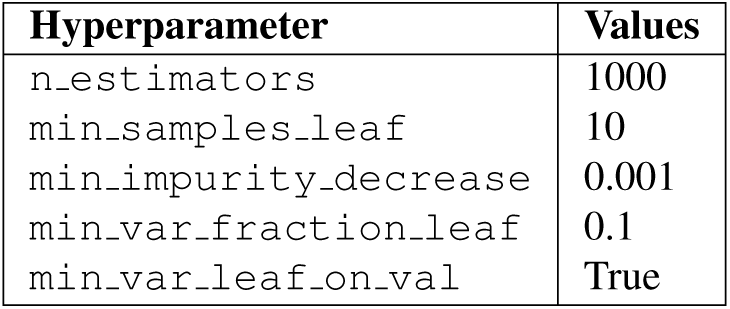
Hyperparameters of the final stage causal forest.

The calculation of the ATE using the AIPW estimator is directly implemented in *EconML* via the method “ate__inference()” when fitting the R-learner for the ITE. This method returns a point estimate for the ATE as well as the corresponding standard error. To get predictions of the ATE for the AUMCdb, we fitted the R-learner also on the AUMCdb dataset.

### Supplement C Assessing the positivity assumption

To ensure that causal effects from observational data are identifiable, three standard assumptions are required (all of which are detailed in Supplement B). Here, we empirically assess the positivity assumption. This assumption states that the propensity score (i.e., the probability of receiving the treatment) for each individual *i* ∈ {*i, . . . , N* }, denoted by *e*(*X_i_*), is bounded by *ɛ < e*(*X_i_*) *<* 1 − *ɛ* with *ɛ >* 0. To evaluate whether this condition is satisfied in our analysis, we plotted the predicted propensity scores, stratified by actual treatment assignment, as shown in Figure S1. We see that the predicted propensity scores are bounded by *ɛ <* 0.006, indicating that no propensity score is excessively close to zero or one. Further, there is a clearly visible overlap between the treated and untreated patients. This suggests that the positivity assumption holds, ensuring that each individual has a non-zero probability of receiving either treatment.

**Figure S1:**
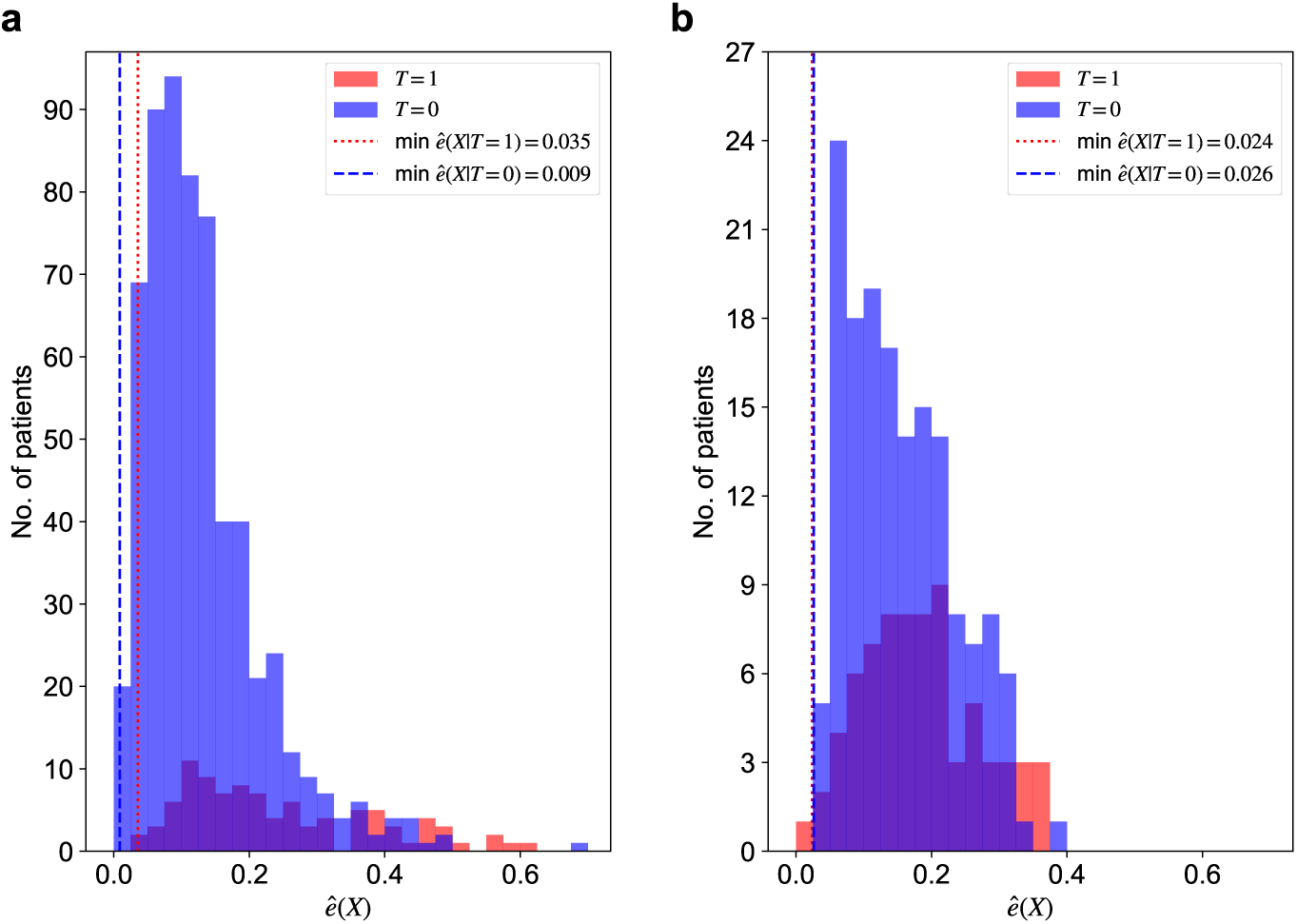
Predicted propensity scores stratified by treatment assignment for **a**, MIMIC-IV and **b**, AUM-Cdb.

### Supplement D Rhythm control

The primary outcome in the main paper is hemodynamic stability since this is the main objective for hemodynamically unstable patients with NOAF in clinical practice. However, rhythm control may be another outcome of clinical relevance. Rhythm control refers to whether sinus rhythm has been restored. This outcome is relevant for hemodynamically unstable patients with NOAF since restoring normal sinus rhythm can help stabilize cardiac function and improve hemodynamics.

#### D.1 Predicted treatment effects

**Figure S2:**
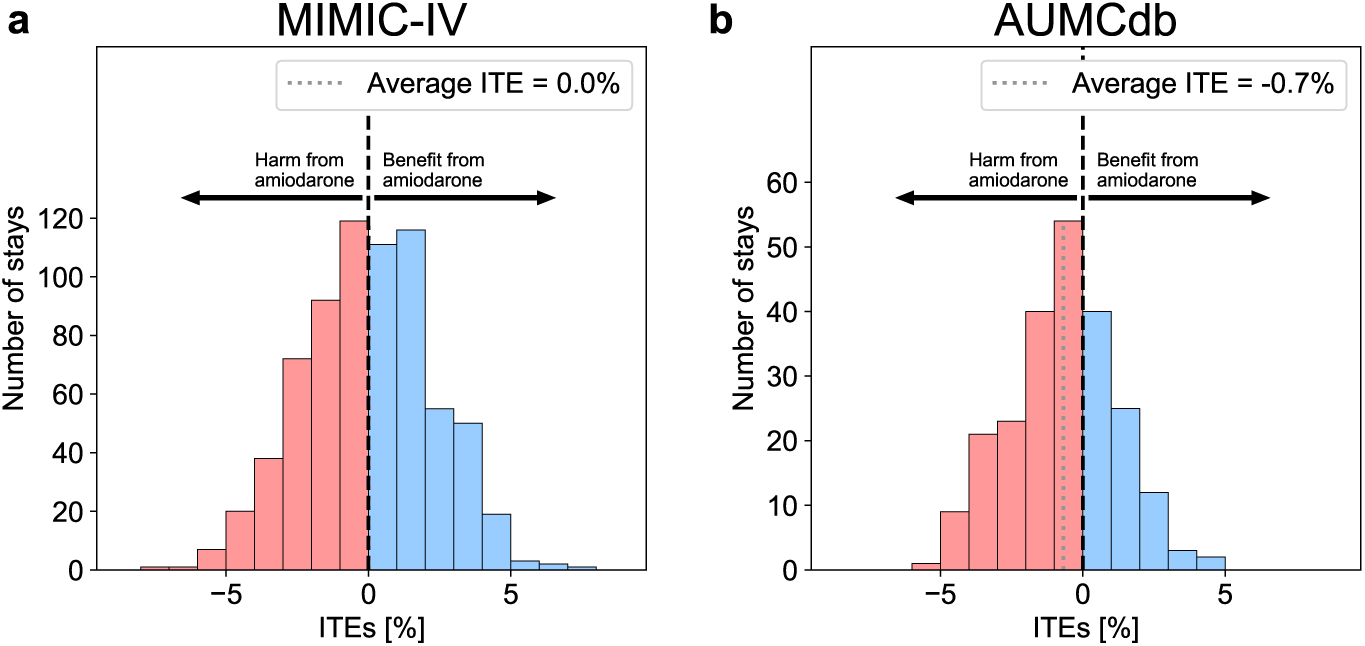
Heterogeneity of predicted ITEs across patients. The histograms show the distribution of predicted ITEs (in %) of amiodarone on restoring sinus rhythm (i.e., rhythm control) and their mean for **a**, MIMIC-IV and **b**, AUMCdb. The distributions of ITEs show that heterogeneity in the effectiveness of amiodarone exists, with values ranging from -3.9% to 3.4% in MIMIC-IV and -5.2% to 4.3% in AUMCdb).

#### D.2 Predictors of treatment effect heterogeneity

**Figure S3:**
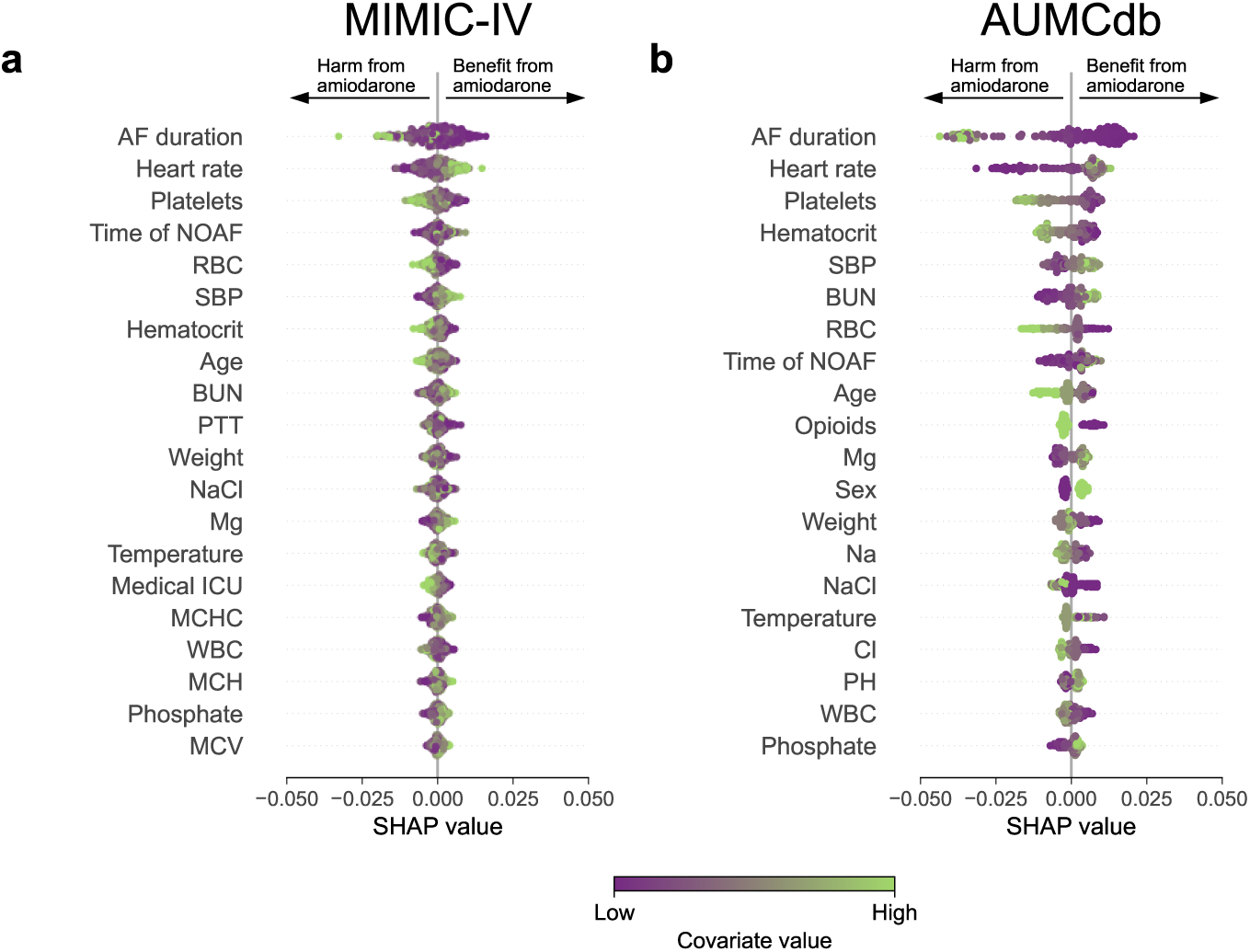
Predictors of treatment effect heterogeneity. The most relevant predictors of treatment effect heterogeneity for **a**, MIMIC-IV and **b**, AUMCdb. The analysis uses SHAP values [45], which helps to explain the contribution of each predictor to the treatment effect predicted by the causal ML model. The predictors are sorted in descending order according to their relevance (top-to-bottom). Each dot in the visualization represents a single ICU stay. The color of the dot indicates the value of the corresponding predictor: green dots represent higher values of the predictor, while purple dots represent lower values. Abbreviations: RBC, red blood cell count; SBP, systolic blood pressure; BUN, blood urea nitrogen; PTT, partial thromboplastin time; NaCl, 0.9% sodium chloride; Mg, magnesium; MCHC, mean corpuscular hemoglobin concentration; WBC, white blood cell count; MCH, mean corpuscular hemoglobin; MCV, mean corpuscular volume; Na, sodium; Cl, chloride.

#### D.3 Personalized treatment rule and validation

**Figure S4:**
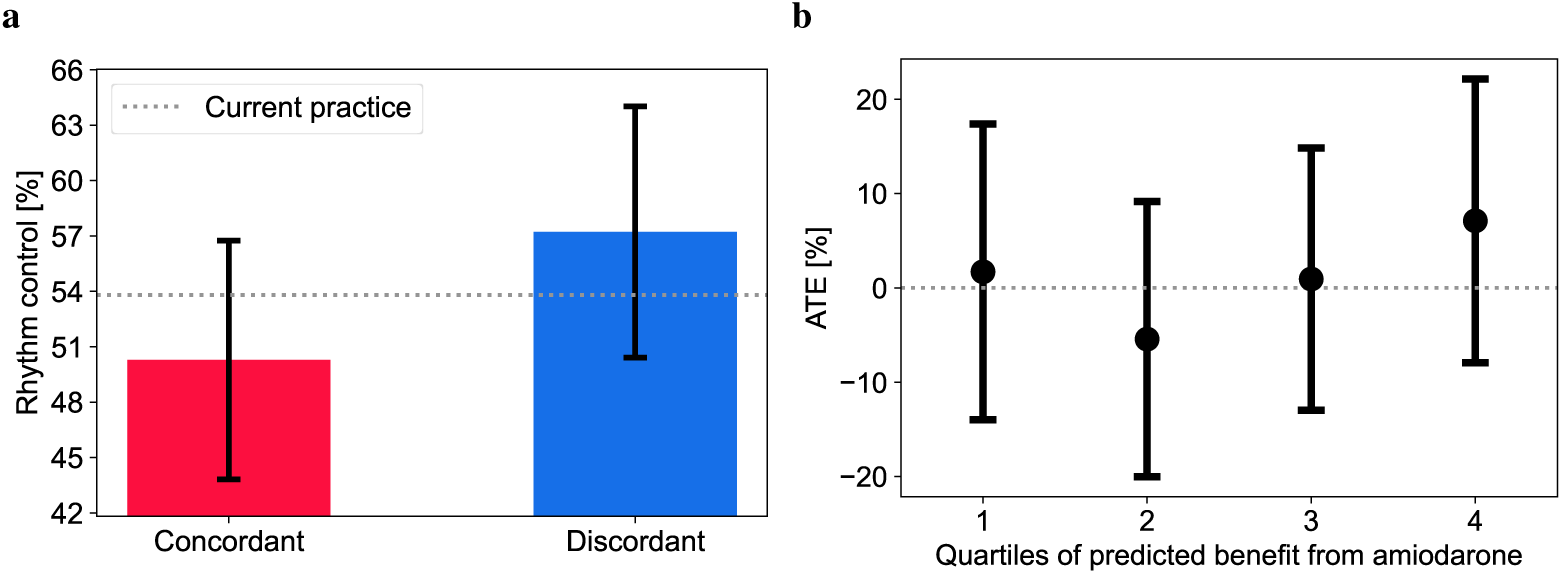
External validation of our causal ML model. **a**, A personalized treatment rule was derived using the predicted ITEs. Average patient outcomes (rhythm control) were compared between patients where our personalized treatment rule was concordant with current practice to patients where it was discordant. The whiskers represent the standard errors (SEs), which were estimated using bootstrapping with 1,000 resamples. As a baseline, the average patient outcome under current practice is shown. **b**, The predicted ITEs for AUMCdb were sorted in descending order and split into quartiles. ATEs were estimated within each quartile. The whiskers represent the SEs.

### Supplement E Rate control

The primary outcome in the main paper is hemodynamic stability since this is the main objective for hemodynamically unstable patients with NOAF in clinical practice. However, besides rhythm, rate control may be another outcome of clinical relevance. Rate control refers to resolving rapid ventricular response. Here, we defined rate control as having a heart rate lower than 110 bpm. This outcome is relevant for hemodynamically unstable patients with NOAF since maintaining a controlled heart rate can reduce the risk of adverse cardiovascular events and can improve hemodynamic stability.

#### E.1 Predicted treatment effects

**Figure S5:**
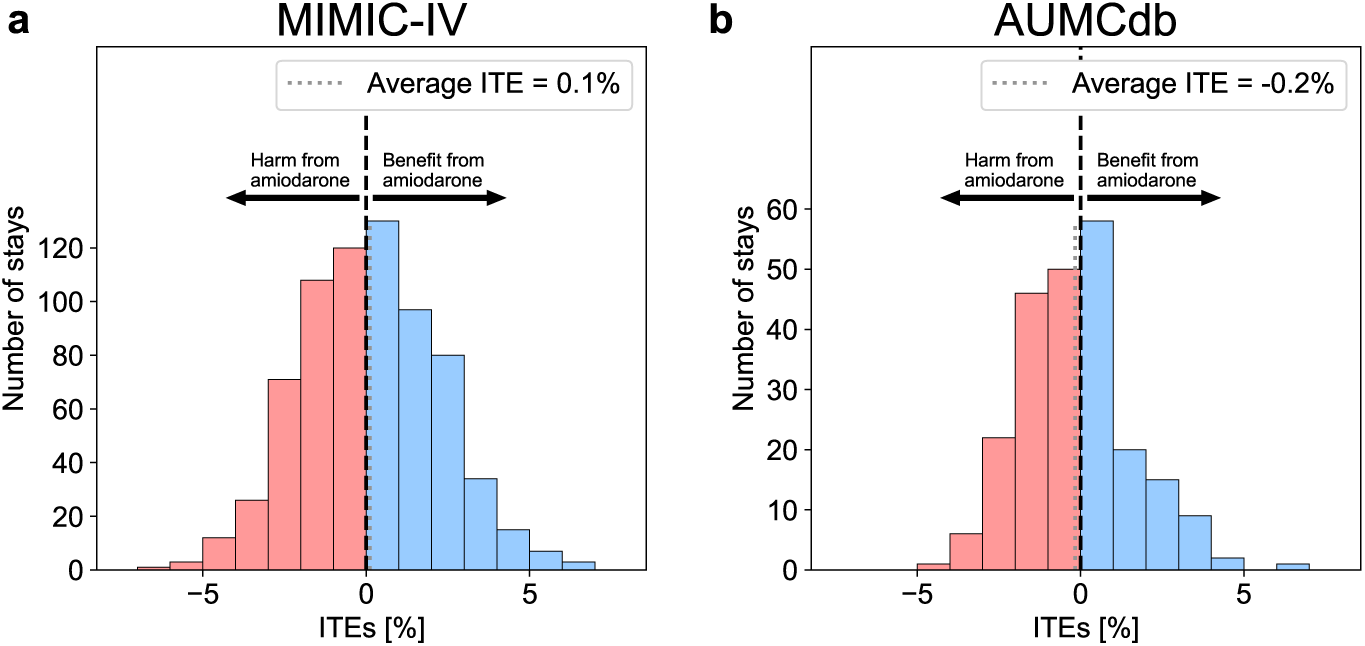
Heterogeneity of predicted ITEs across patients. The histograms show the distribution of predicted ITEs (in %) of amiodarone on rate control and their mean for **a**, MIMIC-IV and **b**, AUMCdb. The distributions of ITEs show that heterogeneity in the effectiveness of amiodarone exists, with values ranging from -2.5% to 2.9% in MIMIC-IV and -4.2% to 6.4% in AUMCdb).

#### E.2 Predictors of treatment effect heterogeneity

**Figure S6:**
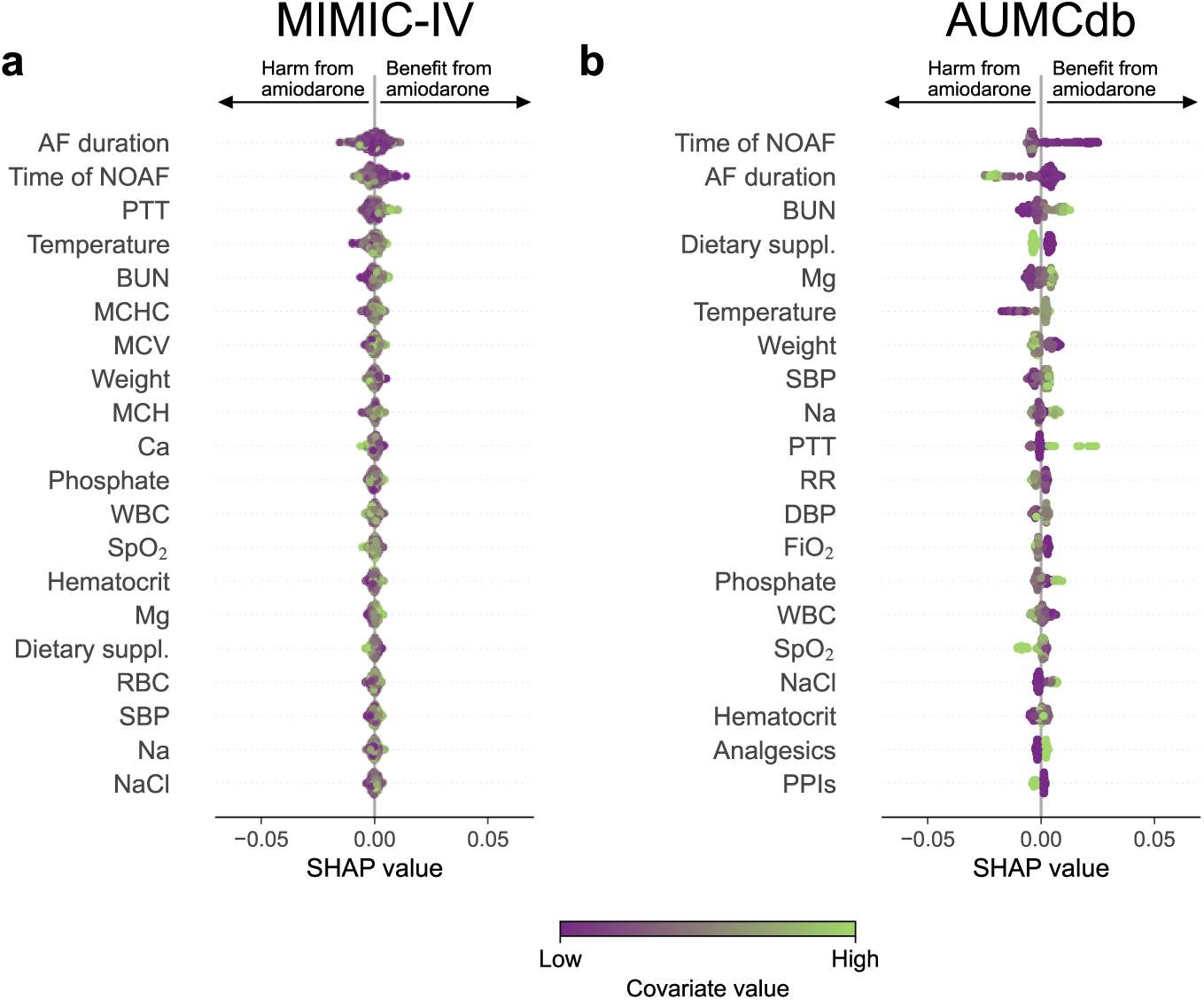
Predictors of treatment effect heterogeneity. The most relevant predictors of treatment effect heterogeneity for **a**, MIMIC-IV and **b**, AUMCdb. The analysis uses SHAP value [45], which helps to explain the contribution of each predictor to the treatment effect predicted by the causal ML model. The predictors are sorted in descending order according to their relevance (top-to-bottom). Each dot in the visualization represents a single ICU stay. The color of the dot indicates the value of the corresponding predictor: green dots represent higher values of the predictor, while purple dots represent lower values. Abbreviations: PTT, partial thromboplastin time; BUN, blood urea nitrogen; MCHC, mean corpuscular hemoglobin concentration; MCV, mean corpuscular volume; Ca, calcium; WBC, white blood cell count; SpO_2_, peripheral oxygen saturation; Mg, magnesium; RBC, red blood cell count; SBP, systolic blood pressure; Na, sodium; NaCl, 0.9% sodium chloride; RR, respiratory rate; DBP, diastolic blood pressure; FiO_2_, fraction of inspired oxygen; PPIs, proton pump inhibitors.

#### E.3 Personalized treatment rule and validation

**Figure S7:**
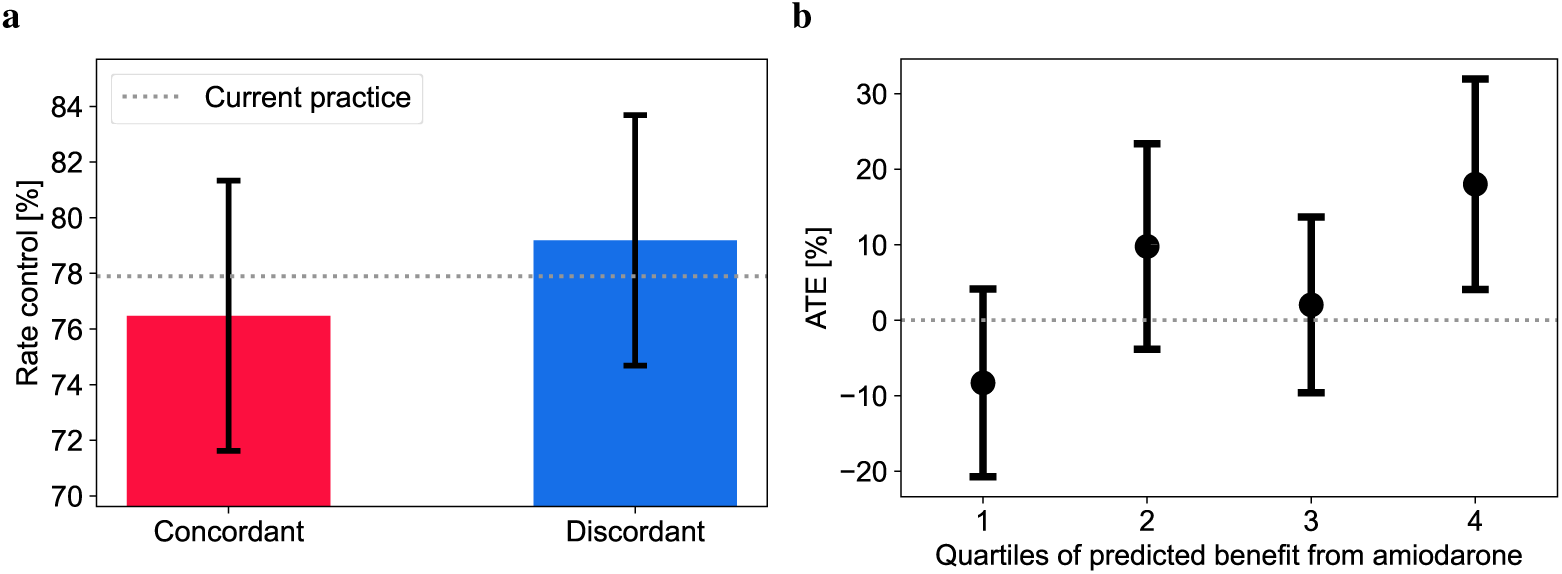
External validation of our causal ML model. **a**, A personalized policy was derived using the predicted ITEs. Average patient outcomes (rhythm control) were compared between patients where our personalized policy was concordant with current practice to patients where it was discordant. The whiskers represent the standard errors (SEs), which were estimated using bootstrapping with 1,000 resamples. As baseline, average patient outcomes under current practice are shown. **b**, The predicted ITEs for AUMCdb were sorted in descending order and split into quartiles. ATEs were estimated within each quartile. The whiskers represent the SEs.

### Supplement F Other ITE estimators

As an additional robustness check, we applied two alternative ITE estimators. In particular, we used the (1) doubly robust (DR) learner [53, 67] with random forests for the treatment assignment and outcome models and (2) the causal forest (CF) [21, 22, 59] as implemented in the *EconML* package. We calculated the Pearson correlation coefficient between the predicted ITEs of our causal ML model (the R-learner) and the two alternative ITE estimators (i.e., DR-learner and CF). We find a high correlation between the predicted ITEs (see Figure S8).

**Figure S8:**
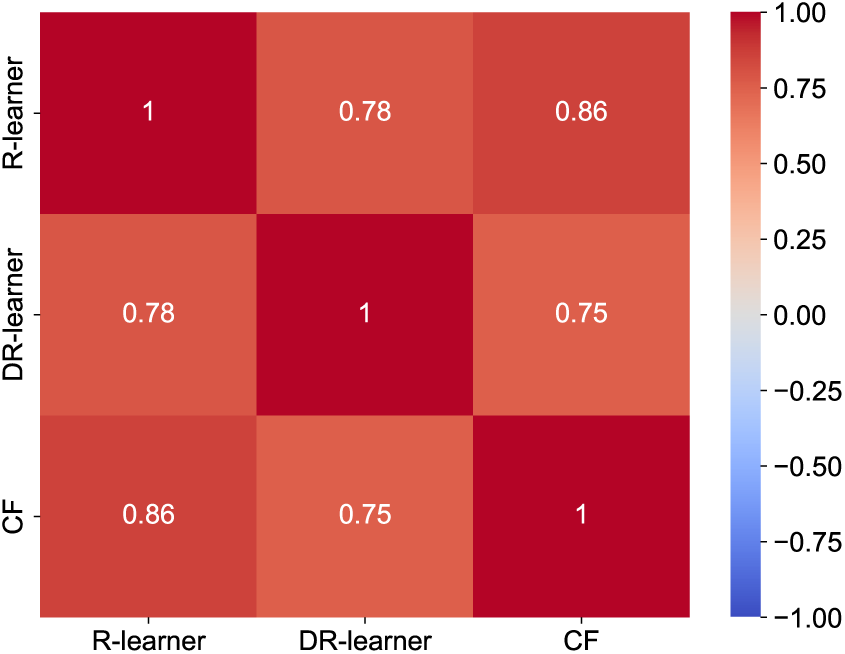
Correlation of predicted ITEs across three different ITE estimators for AUMCdb.

### Supplement G Survival analysis

As an additional robustness check, we conducted a survival analysis. To this end, we defined our outcome as the time until hemodynamic stability is restored. Similar to the main analysis, we assessed the outcome until 12 hours after the end of the treatment window (i.e., 17 hours after time zero). Patient outcomes where hemodynamic stability has not been restored until this point were considered censored. The ITE was then defined as the difference in survival probability under amiodarone treatment vs no treatment. In standard survival analysis, the aim is to maximize survival probability (or the time until the event — typically death — occurs). However, in our setting, we want to minimize the time until hemodynamic stability is restored. Thus, we relabeled the treatment indicator such that a positive ITE also indicates treatment benefit. We performed the analysis using a causal survival forest [61] as implemented in the *grf* package [68].

#### G.1 Predicted treatment effects

Similar to our main analysis, we find that the majority of patients benefit from amiodarone with respect to hemodynamic stability. Specifically, 58.2% of patients in MIMIC-IV and 57.3% of patients in AUMCdb had a positive ITE.

**Figure S9:**
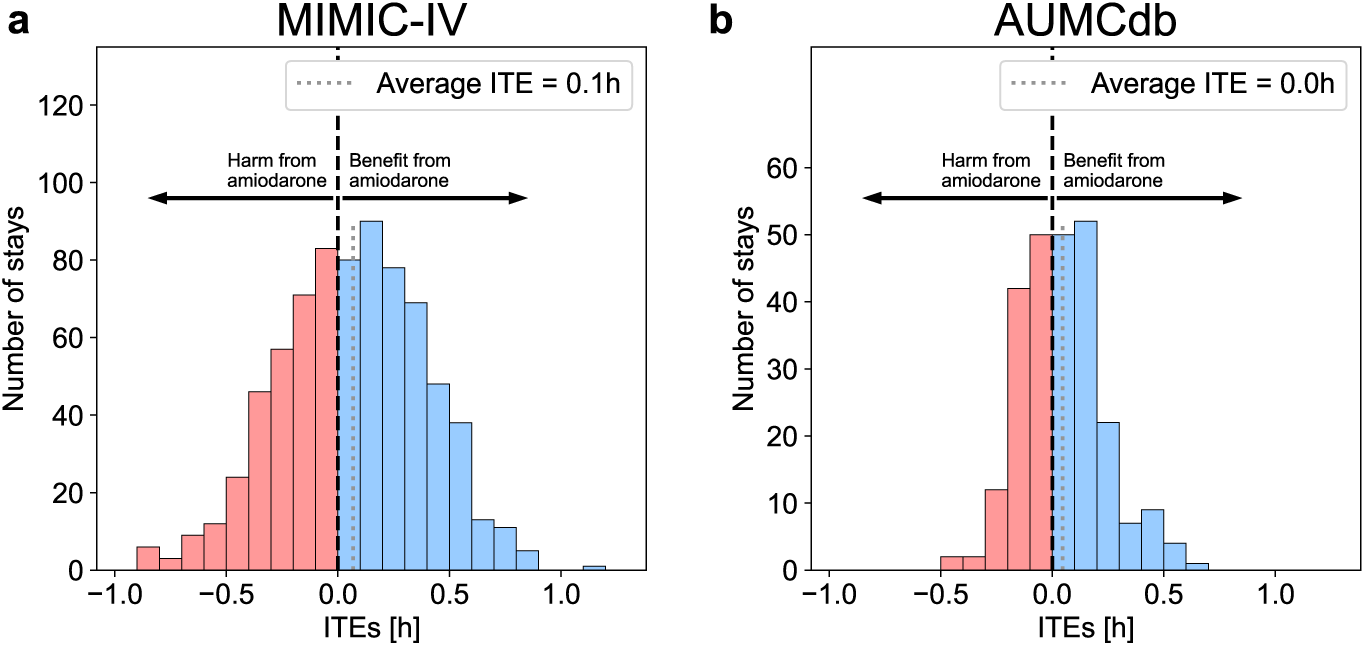
Heterogeneity of predicted ITEs across patients. The histograms show the distribution of predicted ITEs of amiodarone on restoring hemodynamic stability and their mean for **a**, MIMIC-IV and **b**, AUMCdb. The ITE is the difference in probability of restoring hemodynamic stability under amiodarone treatment versus no treatment. The distributions of ITEs show that heterogeneity in the effectiveness of amiodarone exists, further suggesting that a large percentage of patients benefit (58.2% in MIMIC-IV and 57.3% in AUMCdb).

#### G.2 Predictors of treatment effect heterogeneity

We compared the most important predictors of treatment effect heterogeneity of the causal survival forest to those of the main analysis. We find that 11 out of the top-20 covariates are the same. Further, we calculated the Pearson correlation coefficient, which was 0.283(*P* = 0.013).

**Figure S10:**
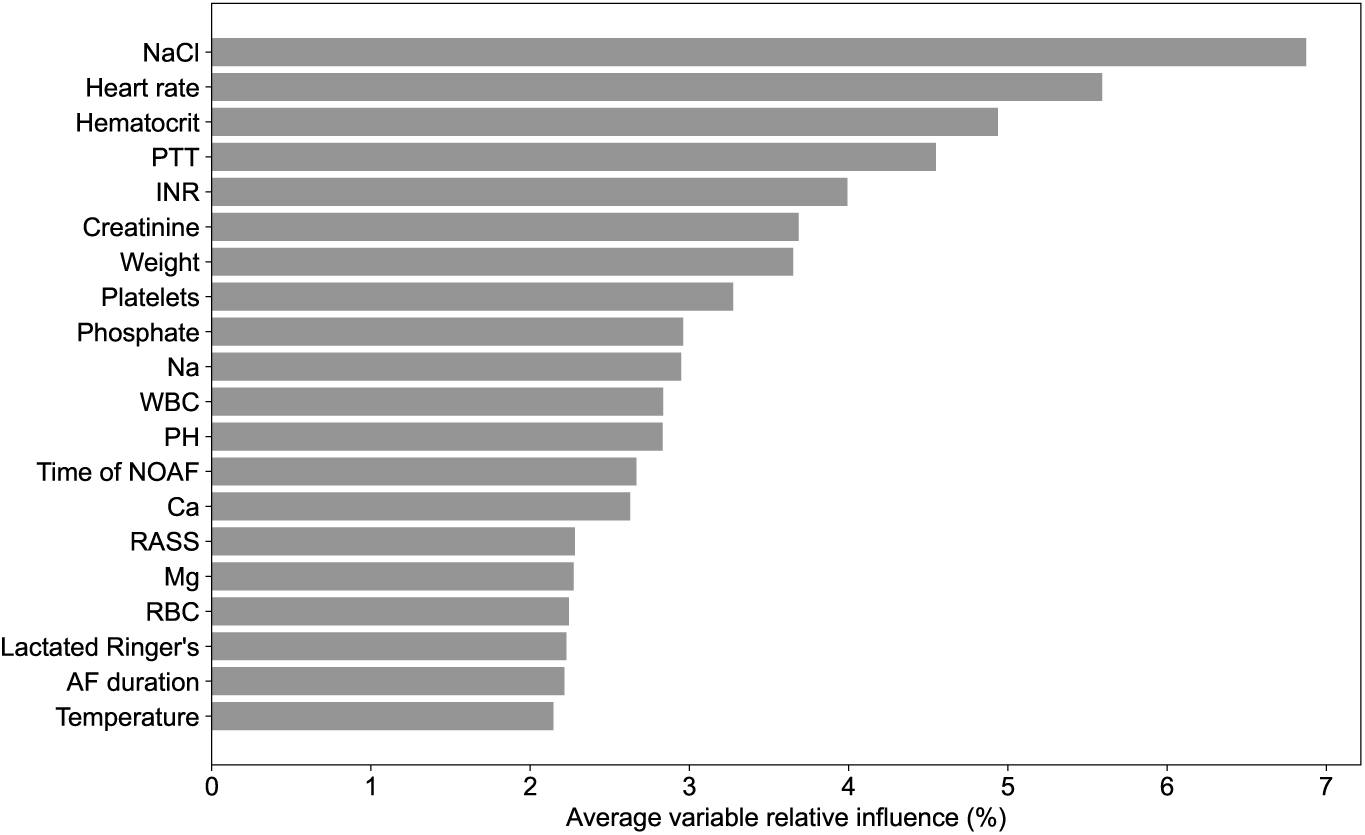
Predictors of treatment effect heterogeneity. The most relevant predictors of treatment effect heterogeneity according to the causal survival forest for **a**, MIMIC-IV and **b**, AUMCdb. The predictors are sorted in descending order according to their relevance (top-to-bottom). Abbreviations: NaCl, 0.9% sodium chloride; PTT, partial thromboplastin time; INR, international normalized ratio; Na, sodium; WBC, white blood cell count; Ca, calcium; RASS, Richmond Agitation-Sedation Scale; Mg, magnesium; RBC, red blood cell count.

#### G.3 Personalized treatment rule and validation

**Figure S11:**
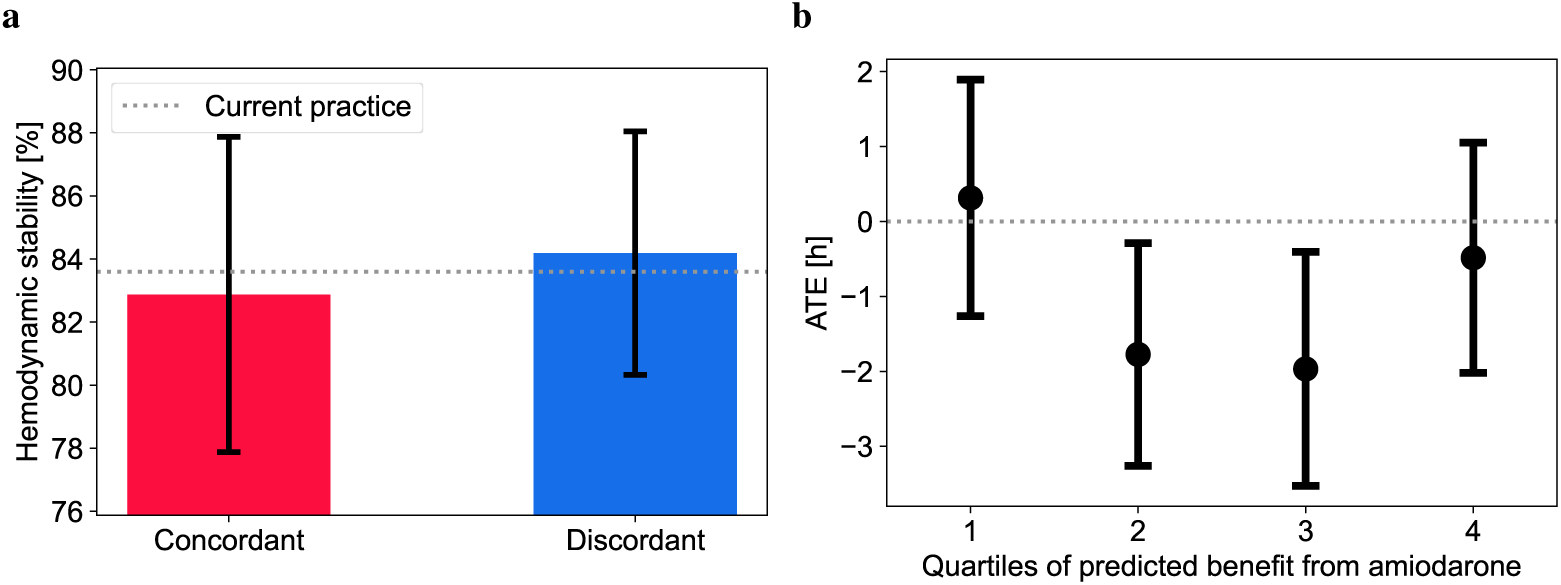
External validation of causal survival forest. **a**, A personalized policy was derived using the predicted ITEs. Average patient outcomes (rhythm control) were compared between patients where our personalized policy was concordant to current practice to patients where it was discordant. The whiskers denote the standard errors (SEs), which were estimated using bootstrapping with 1,000 resamples. As baseline, average patient outcomes under current practice are shown. **b**, The predicted ITEs for AUMCdb were sorted in descending order and split into quartiles. ATEs were estimated within each quartile. The whiskers denote the SEs.

### Supplement H Subgroup analysis

**Table S4:** Characteristics of patients with different treatment effects. Patient populations were split according to their predicted ITE, i.e., into groups that benefit (positive ITE) and groups that are harmed (negative ITE) by amiodarone. We further report the number of patients for which treatment with amiodarone was observed. Dichotomous variables are reported as counts (%), continuous variables as mean (SD). *P* values were calculated using Student’s t-test for continuous variables and the *χ*^2^-test for dichotomous variables. Abbreviations: MAP, mean arterial pressure; SBP, systolic blood pressure; DBP, diastolic blood pressure; RR, respiratory rate; SpO_2_, peripheral oxygen saturation; WBC, white blood cell count; RBC, red blood cell count; IV intravenous; SD, standard deviation.

|  | MIMIC-IV |  |  | AUMCdb |  |  |
| --- | --- | --- | --- | --- | --- | --- |
|  | Positive ITE | Negative ITE | *P* value | Positive ITE | Negative ITE | *P* value |
| ITE | 2.5 (2.5) | -1.4 (1.0) | < 0.001 | 2.6 (2.6) | -1.2 (0.9) | < 0.001 |
| Treated with amiodarone | 39 (5.5%) | 57 (8.1%) | 0.072 | 37 (16.1%) | 36 (15.7%) | 1.0 |
| Age [years] | 73.3 (12.7) | 69.9 (12.0) | < 0.001 | 69.0 (14.7) | 70.7 (12.0) | 0.328 |
| Female | 158 (49.8%) | 194 (50.0%) | 1.0 | 40 (38.8%) | 57 (44.9%) | 0.43 |
| Time of NOAF [hours] | 37.0 (85.5) | 54.4 (71.3) | 0.003 | 44.9 (75.7) | 82.5 (120.1) | 0.006 |
| AF duration [hours] | 6.1 (31.2) | 6.1 (20.1) | 0.999 | 3.1 (6.6) | 10.0 (31.5) | 0.032 |
| MAP [mmHg] | 86.3 (17.6) | 80.1 (13.1) | < 0.001 | 87.3 (22.5) | 80.4 (14.1) | 0.005 |
| SBP [mmHg] | 122.1 (24.4) | 110.8 (18.5) | < 0.001 | 126.7 (36.6) | 118.2 (28.2) | 0.047 |
| DBP [mmHg] | 72.4 (17.7) | 67.1 (13.0) | < 0.001 | 66.4 (18.6) | 63.2 (14.5) | 0.15 |
| Heart rate [bpm] | 130.1 (15.9) | 129.1 (15.4) | 0.379 | 138.2 (20.1) | 135.5 (21.2) | 0.318 |
| RR [breaths/min] | 21.9 (6.2) | 22.6 (6.3) | 0.11 | 19.9 (8.7) | 23.0 (7.6) | 0.006 |
| SpO<sub>2</sub> [%] | 96.1 (2.9) | 95.8 (4.0) | 0.222 | 95.8 (5.5) | 95.5 (3.6) | 0.614 |
| WBC [10<sup>9</sup>/L] | 12.4 (11.5) | 13.5 (8.3) | 0.153 | 12.9 (8.7) | 15.2 (6.6) | 0.021 |
| RBC [10<sup>12</sup>/L] | 3.4 (0.6) | 3.2 (0.6) | < 0.001 | 3.4 (0.5) | 3.3 (0.4) | 0.231 |
| IV norepinephrine [μg/kg/min] | 0.046 (0.124) | 0.031 (0.085) | 0.07 | 0.133 (0.215) | 0.102 (0.179) | 0.238 |
| Mechanical ventilation | 105 (33.1%) | 133 (34.3%) | 0.808 | 68 (66.0%) | 75 (59.1%) | 0.344 |

### Supplement I Refutation checks

We applied three different refutation checks to assess the robustness of our causal ML model. To this end, we repeated the analyses with a random treatment, a random outcome, and by adding random covariates to the actual covariates [62]. In the first two cases, the ITEs should disappear and in the last case, the ITEs should remain robust. This mostly aligns with our observations, as shown in Figure S12.

**Figure S12:**
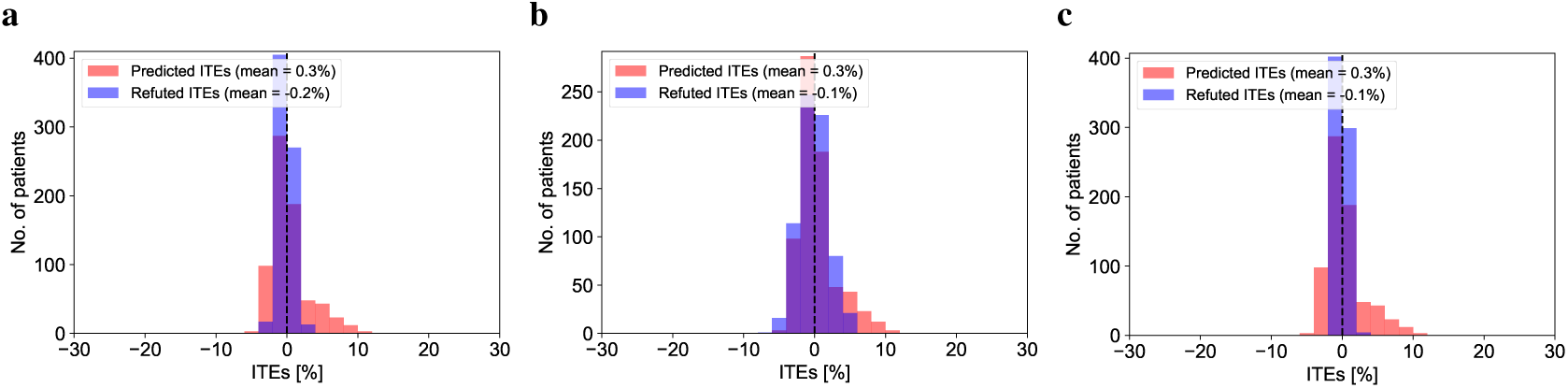
Refutation checks where we proceed as follows: **a**, replace the true treatment assignments by random treatments; **b**, replace the true outcomes by random outcomes; and **c**, add random covariates to the existing, real covariates. Since we used the MIMIC-IV dataset for developing the causal ML model, we only show the results for this dataset.

### Supplement J Calibration

We assessed the calibration of our treatment assignment and outcome models, which were implemented using random forests [58]. Calibration refers to the degree to which the predicted probabilities from a model agree with the actual observed frequencies of the outcome. Results are shown in Figure S13.

**Figure S13:**
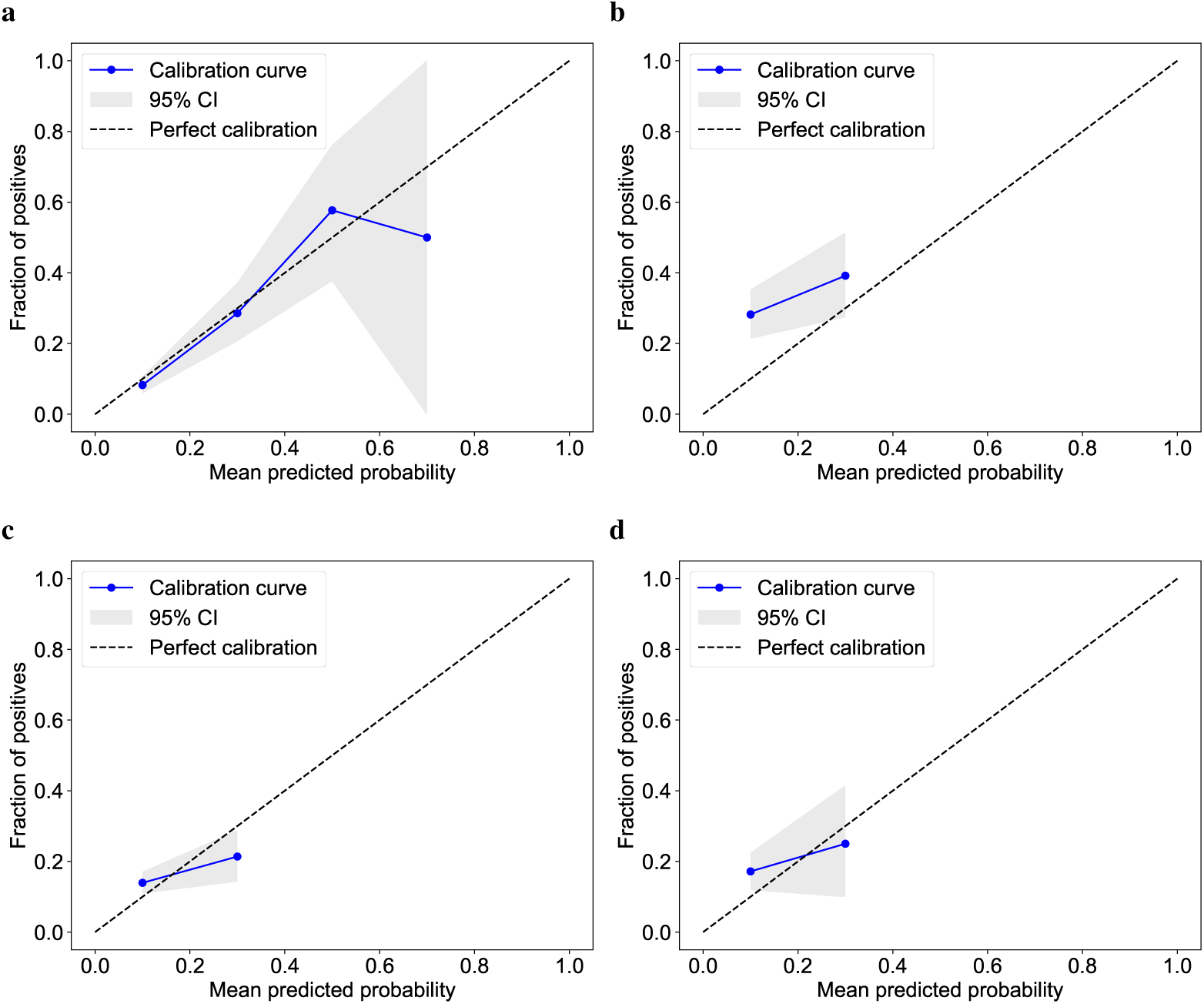
Assessment of model calibration for the treatment assignment model for MIMIC-IV (**a**) and AUMCdb (**b**) and for the outcome model for MIMIC-IV (**c**) and AUMCdb (**d**).

## Notes

### Competing Interest Statement

The authors have declared no competing interest.

### Author Declarations

The source data was available before the initiation of the study and can be accessed after successful application via https://physionet.org/content/mimiciv/2.0/ and https://amsterdammedicaldatascience.nl/#amsterdamumcdb.

